# Influenza vaccination and single cell multiomics reveal sex dimorphic immune imprints of prior mild COVID-19

**DOI:** 10.1101/2022.02.17.22271138

**Authors:** Rachel Sparks, William W. Lau, Can Liu, Kyu Lee Han, Kiera L. Vrindten, Guangping Sun, Milann Cox, Sarah F. Andrews, Neha Bansal, Laura E. Failla, Jody Manischewitz, Gabrielle Grubbs, Lisa R. King, Galina Koroleva, Stephanie Leimenstoll, LaQuita Snow, OP11 Clinical Staff, Jinguo Chen, Juanjie Tang, Amrita Mukherjee, Brian A. Sellers, Richard Apps, Adrian B. McDermott, Andrew J. Martins, Evan M. Bloch, Hana Golding, Surender Khurana, John S. Tsang

## Abstract

Viral infections can have profound and durable functional impacts on the immune system. There is an urgent need to characterize the long-term immune effects of SARS-CoV-2 infection given the persistence of symptoms in some individuals and the continued threat of novel variants. Here we use systems immunology, including longitudinal multimodal single cell analysis (surface proteins, transcriptome, and V(D)J sequences) from 33 previously healthy individuals after recovery from mild, non-hospitalized COVID-19 and 40 age- and sex-matched healthy controls with no history of COVID-19 to comparatively assess the post-infection immune status (mean: 151 days after diagnosis) and subsequent innate and adaptive responses to seasonal influenza vaccination. Identification of both sex-specific and -independent temporally stable changes, including signatures of T-cell activation and repression of innate defense/immune receptor genes (e.g., Toll-like receptors) in monocytes, suggest that mild COVID-19 can establish new post-recovery immunological set-points. COVID-19-recovered males had higher innate, influenza-specific plasmablast, and antibody responses after vaccination compared to healthy males and COVID-19-recovered females, partly attributable to elevated pre-vaccination frequencies of a GPR56 expressing CD8+ T-cell subset in male recoverees that are “poised” to produce higher levels of IFNγ upon inflammatory stimulation. Intriguingly, by day 1 post-vaccination in COVID-19-recovered subjects, the expression of the repressed genes in monocytes increased and moved towards the pre-vaccination baseline of healthy controls, suggesting that the acute inflammation induced by vaccination could partly reset the immune states established by mild COVID-19. Our study reveals sex-dimorphic immune imprints and *in vivo* functional impacts of mild COVID-19 in humans, suggesting that prior COVID-19, and possibly respiratory viral infections in general, could change future responses to vaccination and in turn, vaccines could help reset the immune system after COVID-19, both in an antigen-agnostic manner.

## Introduction

Infection with SARS-CoV-2 can result in persistent clinical sequelae for months after initial infection, both in those requiring hospitalization and those with mild disease^1^. While the spectrum of clinical manifestations associated with post-acute COVID-19 syndrome (a.k.a “long COVID”) is expanding, understanding the molecular and cellular immunological changes associated with recovery from SARS-CoV-2 infection is lacking, particularly in those with less severe, non-hospitalized disease, the population that constitutes the majority of COVID-19 recoverees. Important questions include how “homeostatic”/baseline immune states may have been altered by the infection, and whether any alterations may affect responses to future challenges (e.g., infection or vaccination). Examples of long-term immunological effects of viral infection and innate “training” effects of vaccination^2^ have previously been described, e.g., following natural measles infection there is marked reduction in humoral immunity and increased susceptibility to various non-measles infections for months to years^3, 4^. A better understanding of whether even mild SARS-CoV-2 infection could result in persistent immunological changes that may affect future immune responses has important public health implications given the large number of infected individuals in the world (more than 500 million global cases as of July, 2022)^5, 6^. These natural viral infections also provide an unprecedented opportunity to assess how a respiratory infection may change the immune status in humans. We thus enrolled and comparatively analyzed using systems immunology, including multimodal single cell approaches, healthy, non-obese individuals who: 1) recovered from non-hospitalized, mild cases of COVID-19, and 2) age- and sex-matched controls who never had COVID-19. In addition to assessing the post-COVID-19 immunological states, we utilized seasonal influenza vaccination to evaluate the immune responses of these two populations at the serological, transcriptional, proteomic, and cellular levels. These assessments together help reveal basic principles regarding what happens to the immune system after two well-defined immunological encounters in humans: mild COVID-19 as a natural infectious perturbation and influenza vaccination as a controlled and timed intervention.

## Results

Individuals with prior symptomatic SARS-CoV-2 infection (n=31; diagnosed by nasal PCR test) or asymptomatic infection (n=2; by antibody test, see Methods) during the first wave of COVID-19 in 2020, and age- and sex-matched healthy controls (HC; n=40) with no history of COVID-19 (and negative by antibody test) were recruited from the community in Fall 2020 and followed longitudinally (Fig. 1a, see Methods). For those with a history of COVID-19, the average time since diagnosis was 151 days (Extended Data Fig. 1a; Extended Data Table 1). In addition, two individuals had asymptomatic COVID-19 infection, defined as positive for antibodies against SARS-CoV-2 with no history of symptoms or positive nasal PCR test. All COVID-19-recovered (COVR) individuals had clinically mild illness during acute disease that did not require hospitalization (self-reported average length of illness: 16.5 days), nor did they have any major medical comorbidities, including infection at the time of enrollment, obesity (BMI > 30) or autoimmune disease (Fig. 1b). None of the participants were enrolled in COVID-19 vaccine trials, nor did they receive recent vaccination of any kind before administration of the seasonal influenza vaccine in this study. A small number of individuals continued to have mild self-reported sequelae from their illness at study enrollment (3 males and 8 females), the most common being loss of taste and/or smell (Extended Data Table 1). Females tended to be more likely to have sequelae (Fisher’s exact test p = 0.09 for all subjects, p = 0.03 for those < 65 years of age), at a rate similar to that reported in other larger studies^7^.

**Figure 1.**
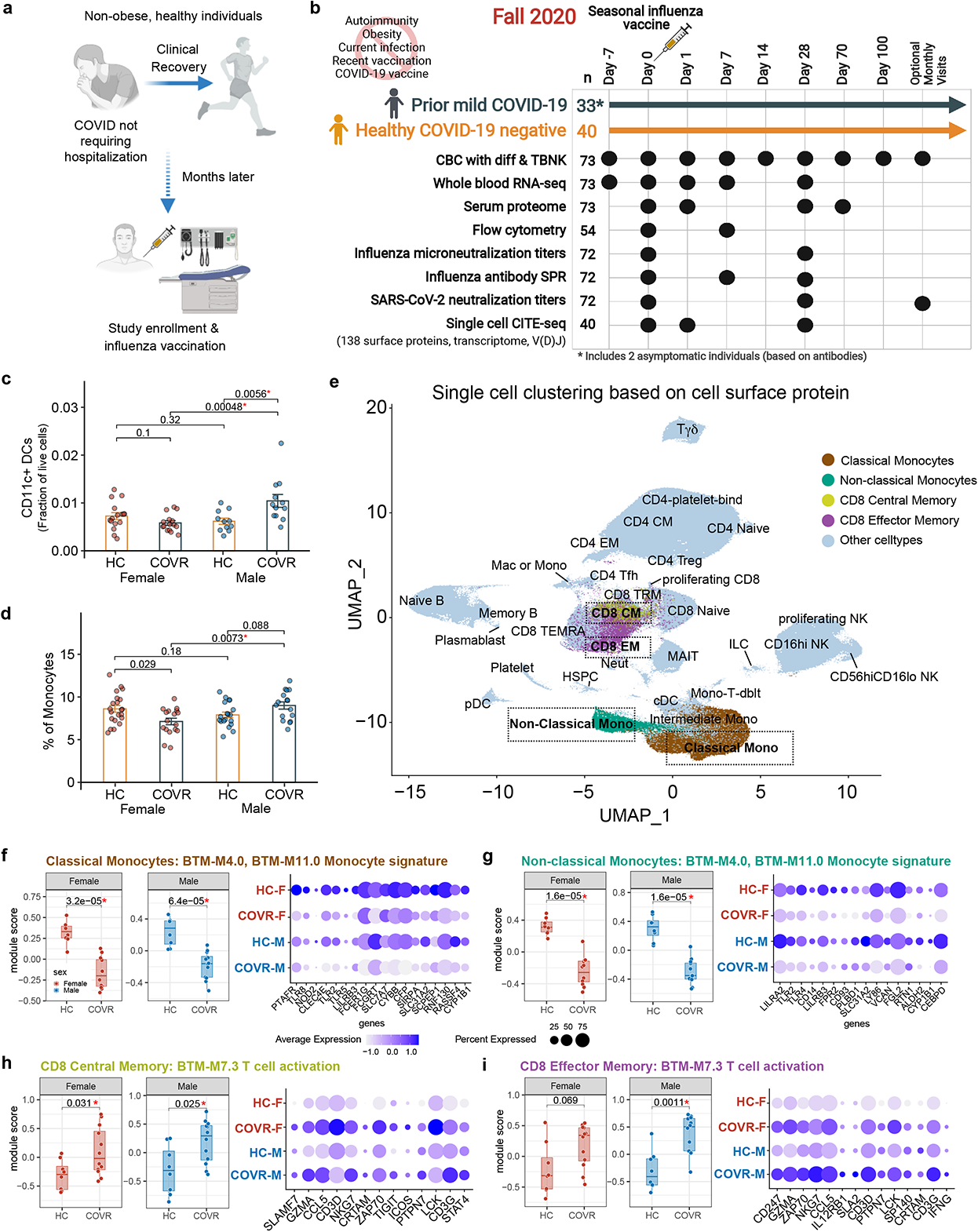
Study overview and evaluation of baseline (before influenza vaccination) molecular and cellular differences between COVID-19-recovered subjects and matching controls. **a**, Schematic showing the study concept and design. **b**, Data generated at each timepoint in the study. Both COVID-19-recovered (COVR) subjects and healthy controls (HC) were enrolled at seven days before vaccination [day -7 (pre7)] and sampled longitudinally at the indicated timepoints relative to the day of influenza vaccination. The total number of subjects assayed for each data type is indicated next to the assay. CBC with diff & TBNK = complete blood count with differential and T- and B-lymphocyte and Natural Killer cell phenotyping. SPR = Surface plasmon resonance **c**, Bar plots comparing the proportion of CD11c+ dendritic cells (DCs; as the fraction of live cells from flow cytometry; y-axis) between COVR females (COVR-F; n = 15), HC females (HC-F; n = 16), COVR males (COVR-M; n = 12), and HC males (HC-M; n = 11) at day 0 (D0). Significance of differences is determined by two-tailed Wilcoxon test. **d**, Similar to (**c**) but for the percentage of monocytes in peripheral blood (measured by the CBC; y-axis) between COVR-F (n = 17), COVR-M (n = 16), HC-F (n = 21), and HC-M (n = 19) at baseline (average of pre7 and D0). Significance of differences is determined by two-tailed Wilcoxon test. **e**, UMAP visualization of the CITE-seq single cell data showing clustering of cells based on cell surface protein markers (632,100 single cells from all timepoints with CITE-seq data: days 0, 1, 28). Cell-specific clusters are labeled. Cell populations of interest are boxed. Colored and boxed cell clusters are further explored in (**f-i**). **f**, (left) Box plots comparing the Gene Set Variation Analysis (GSVA) module scores [using the union of the leading-edge genes (LEGs) from monocyte gene sets BTM M4.0 and M11.0; see Methods; y-axis] between HC-F (n = 8) and COVR-F (n = 12) (left box) and HC-M (n = 8) and COVR-M (n = 12) (right box) using the CITE-seq classical monocyte pseudobulk expression data at D0. Each point represents a subject. Significance determined by two-tail Wilcoxon test. (right) Bubble plot showing the average gene expression of selected LEGs, including those in the Gene Ontology (GO) pattern recognition receptor activity and immune receptor activity gene sets. **g**, Similar to (**f**) but showing the non-classical monocyte population at D0. **h**, (left) Box plots comparing the T-cell activation gene set (BTM-M7.3) GSVA module scores (y-axis) between HC-F (n = 8) and COVR-F (n = 12) (left box) and HC-M (n = 8) and COVR-M (n = 12) (right box) for the CITE-seq CD8+ T-cell central memory at D0. Significance determined by two-tailed Wilcoxon test. (right) Bubble plot showing the average gene expression of the selected LEGs shared by male and female from the gene set enrichment analysis (see Methods) **i**, Similar to (**h**) but showing the CD8+ T-cell effector memory population at D0.

### Prior mild COVID-19 is associated with stable sex-specific molecular and cellular differences

Longitudinal multi-omics profiling was performed using whole blood transcriptomics, single cell analysis of 138 surface proteins, transcriptome, and V(D)J sequences via CITE-seq (Cellular Indexing of Transcriptomes and Epitopes by Sequencing^8^), serum protein profiling, antibody characterization, peripheral blood immune cell frequencies with hematological parameters from a complete blood count (CBC) as well as clinical and research flow cytometry covering major immune cell lineages and subsets (Fig. 1b, Supplementary Information Fig. 1, Supplementary Information Tables 1 and 2). We first assessed baseline, pre-vaccination differences between the COVR individuals and the age- and sex-matched HCs. As sex-dependent acute immune responses to COVID-19 have been reported^9–17^, our analyses explicitly searched for sex-dependent signatures associated with prior mild COVID-19. Immunological resolution following infection may unfold over time even after symptoms subside, and there were indeed parameters that showed evidence of continued evolution in our cohort—defined as those that were correlated with time since COVID-19 diagnosis (TSD; Extended Data Table 2, see Methods; asymptomatic individuals were not included in TSD analyses), such as SARS-CoV-2 neutralizing antibody titers^18^ (Extended Data Fig. 1b). However, we were primarily interested in understanding persistent post-COVID-19 immune imprints, and thus our analyses focused on temporally stable immune states associated with prior mild COVID-19, defined as those that were *not* significantly associated with TSD.

We evaluated differences between 1) COVR females (COVR-F) vs. HC females (HC-F); 2) COVR males (COVR-M) vs. HC males (HC-M); and 3) COVR-M vs. COVR-F after accounting for male-female differences in HCs (herein referred to as “sex differences”; Extended Data Tables 3 and 4). We noted that the frequencies of myeloid cells such as monocytes and conventional/myeloid dendritic cells (cDCs) tend to be higher in COVR-M than HC-M and/or COVR-F, differences that are not associated with TSD (Fig. 1c, d; Extended Data Fig. 1c, d). Whole blood transcriptomics data also revealed sex-dependent signatures associated with prior COVID-19 (Extended Data Fig. 1e; e.g., the monocyte-related M11.0 and M4.0 from the blood transcriptional module [BTM] collection), including sex differences in metabolic transcriptional signatures such as oxidative phosphorylation. However, blood transcriptomic signatures can be driven by both cell composition and cell intrinsic transcriptional differences. Indeed, the innate immune, metabolic, and T-cell-related signatures are driven, at least in part, by the increased circulating monocyte and correspondingly lower T-cell frequencies in COVR-M (Fig. 1d and Extended Data Fig. 1f) as these enrichment signals became statistically insignificant when monocyte frequencies were included as a covariate in the model (data not shown).

To assess transcriptional signatures independent of cell frequencies, we used CITE-seq data to examine cell type-dependent contributions underlying the whole blood transcriptomic signatures. We clustered single cells and annotated the resulting clusters using surface protein expression profiles (see Methods) to reveal major immune cell lineages and subsets (Fig. 1e). Cell type-specific transcriptional analysis pointed to both sex-dependent and -independent differences between COVR and HCs (Extended Data Table 5). Among the enriched gene sets in whole blood transcriptomic analysis from above (Extended Data Fig. 1e), but now free of confounding due to the cell frequency differences in monocytes and other cell lineages, the BTM M11.0/4.0 gene sets exhibit depressed expression in both classical and non-classical monocytes in COVR relative to HCs in both sexes, while the converse is true for genes in the T-cell activation signature (BTM M7.3) in both CD8+ central memory and effector memory (EM) T-cells (Fig. 1f-i; Extended Data Fig. 1g; Extended Data Table 6). The T-cell activation signature in CD8+ EMs was particularly pronounced in COVR-M (Fig. 1i). The genes with lower expression in the COVR monocytes driving the above enrichment [i.e., the so-called “leading edge genes” (LEGs)] include numerous surface receptors, such as those encoding pattern recognition receptors (TLR2, TLR4, and TLR8), the peptidoglycan recognizing receptor NOD2, the high affinity IgE FC receptor FCER1G, and C-type lectin receptor CLEC4A (Fig. 1f, g). Both the monocyte depression and T-cell activation signatures are predominantly not associated with TSD in both males and females (Extended Data Fig. 1h), indicating that they are persistent, temporally stable features established by the acute infection.

The T-cell activation signature identified in the single cell data likely emerged during and persisted after acute COVID-19^19^, but this was less clear for the monocyte depression signature. We thus asked whether the monocyte signature could be linked to gene expression changes seen in acute SARS-CoV-2 infection. Using a previously published CITE-seq dataset we generated from a hospitalized, predominantly older and male-biased severe COVID-19 cohort from Italy^19^, we noted that within the classical monocytes, the average expression of the LEGs noted above was significantly lower in acute COVID-19 patients than healthy controls and negatively associated with disease severity (Extended Data Fig. 1i). Thus, this depressed monocyte transcriptional signature could have originated from and stably persisted since the early acute response to the infection. Several studies have also reported the emergence of several (potentially overlapping) types of altered monocytes in acute COVID-19, including those with lower antigen presentation, depressed NF-kB/inflammation, or myeloid-derived suppressor cell (MDSC)-like phenotypes^19–24^. However, none of these monocyte phenotypes were found to be significantly different in the monocytes of COVR compared to HCs in our cohort at baseline before influenza vaccination (Supplementary Information Figure 2), suggesting that the depressed monocyte gene signature involving pattern recognition and immune receptor genes observed here is unique and distinct from those identified in acute disease. Together, our findings suggest that even mild, non-hospitalized SARS-CoV-2 infections may establish new, temporally stable, sex-dependent immunological imprints.

To assess whether other natural respiratory viral infections may leave similar unresolved sex-specific “immune states”, we used a published blood transcriptomic dataset profiling two independent cohorts of patients with confirmed community influenza A (predominantly pandemic H1N1) infection during two influenza infection seasons (2009-2010 and 2010-2011, respectively)^25^. By comparing the blood transcriptomic profiles before and after the season (I.e., before infection and post-recovery), we found robust changes consistent between the two cohorts/seasons in males, but the changes across the two cohorts were largely not correlated in females (Extended Data Fig. 2a,b; Extended Data Table 7). The genes with increased expression in males after recovery from natural influenza infection (those consistent between the two cohorts/seasons) were also enriched for genes more highly expressed in COVR-M compared to COVR-F in our cohort (after accounting for the expected sex differences present in healthy subjects; Extended Data Fig. 2c). In addition, the genes with lower expression after influenza infection and recovery in males were enriched for the depressed classical and non-classical monocyte signatures above including TLR5 (Fig. 1f,g; FDR<0.05 Fisher’s Exact Test). These observations provide independent support that exposure to a respiratory viral pathogen can lead to persistent immunological imprints detectable in blood, even in healthy individuals with mild disease, although in general some of these imprints are likely pathogen-dependent.

### Prior mild COVID-19 is associated with heightened innate and adaptive responses to influenza vaccination in males

We next asked whether prior COVID-19 may impact an individual’s response to non-SARS-CoV-2 immunological challenges. The seasonal influenza quadrivalent vaccine was administered to study participants, who were subsequently followed longitudinally for up to 100 days, including days 1, 7, and 28, to evaluate the immune response to the vaccine at the serological, molecular, and cellular levels (Fig. 1a, 1b, 2a). This vaccine was selected in part due to its public health importance: the 2020-21 influenza season was approaching at the start of our study and it was not clear whether prior COVID-19 infection would impact responses to the influenza vaccine. In addition, the cellular and molecular responses to seasonal influenza vaccination and the associated kinetics have been well characterized in healthy adults, including early innate/inflammatory and interferon (IFN) responses on day 1 (D1) after vaccination and a strong but transient plasmablast peak around day 7 (D7) culminating in the generation of influenza-specific antibodies^26–29^. Thus, influenza vaccination provides an excellent perturbation to elicit coordinated immune activity that can be sampled at specific times after vaccination to probe the potential functional impacts of prior mild SARS-CoV-2 infection.

**Figure 2.**
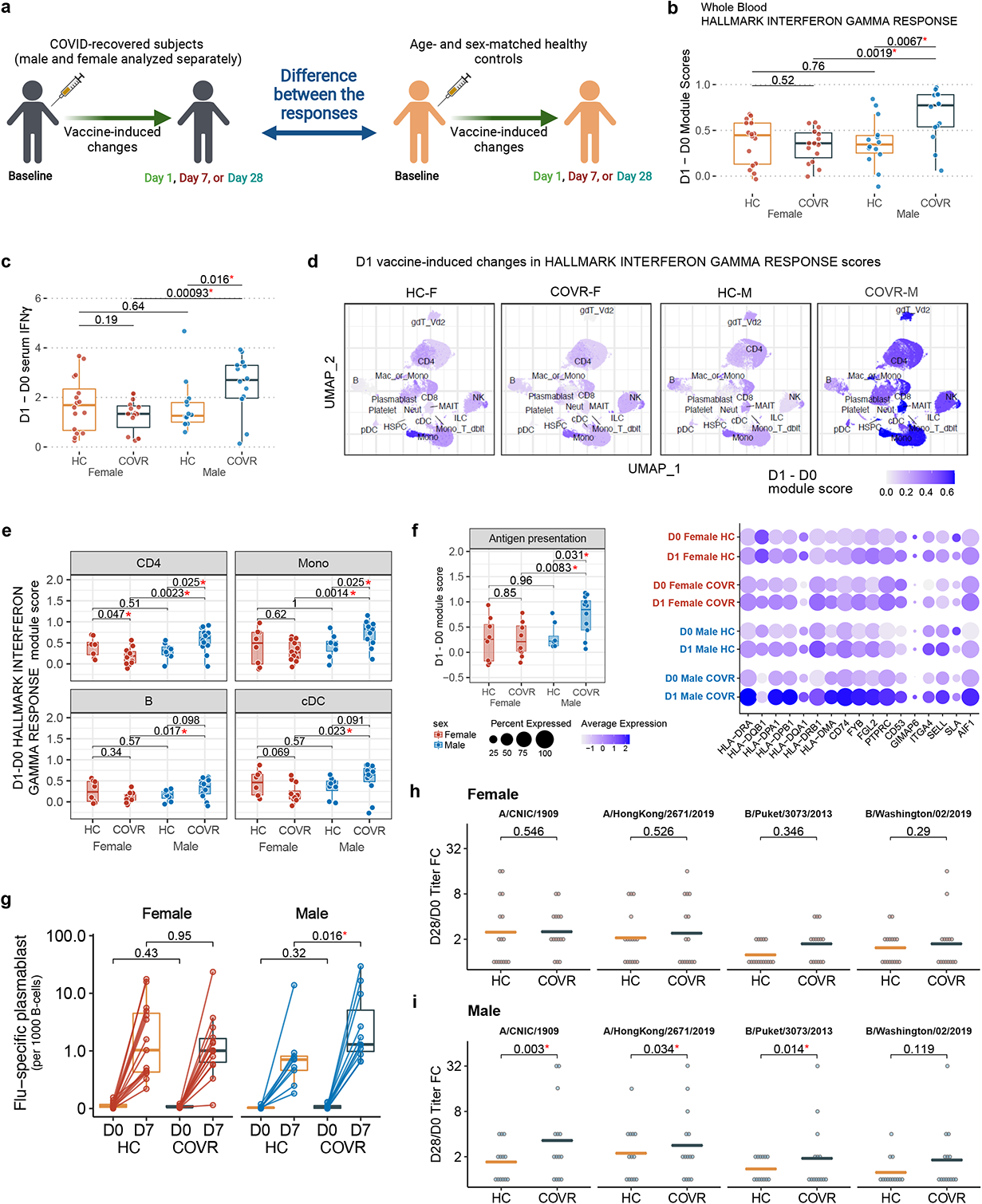
Sex-specific molecular, cellular, and humoral response differences to influenza vaccination between COVID-19-recovered individuals and matching controls. **a**, Schematic of the sex-specific comparisons of influenza-vaccine induced changes from baseline (pre-vaccination) at timepoints post vaccination (D1, D7, and D28) between COVID-19-recovered (COVR) subjects and healthy control (HC) subjects. Subsequent analyses limited to subjects under 65 years of age who received the Flucelvax quadrivalent seasonal influenza vaccine. **b**, Box plots showing the difference between D1 and day 0 (D0) in the whole blood Hallmark Interferon Gamma (IFNγ) Response module score for COVR females (COVR-F; n=15), COVR males (COVR-M; n=14), and HC females (HC-F; n=16), and HC males (HC-M; n=14). Significance of differences between groups is determined by two-tailed Wilcoxon test. **c**, Box plots of the D1 response (D1 – D0) of serum IFNγ protein level (from the OLINK platform) for the subjects in shown in (**b**). Significance of differences between groups is determined by two-tailed Wilcoxon test. **d**, Surface protein-based UMAP visualization of single cell CITE-seq data showing the D1 response (D1 – D0) in the Hallmark IFNγ Response module score within each cell cluster/subset for HC-F (n = 8), COVR-F (n=12), and HC-M (n=8), COVR-M (n=12). Coarser level cell types are labeled. Darker color indicates a greater difference between D1 and D0 for the indicated cell type. **e**, Boxplots comparing the D1 response (D1 – D0) in the Hallmark IFNγ Response module score (y-axis) for the indicated cell types from the CITE-seq pseudobulk expression for HC-F (n = 8), COVR-F (n = 12), HC-M (n = 8) and COVR-M (n = 12). Significance of group difference is determined by two-tailed Wilcoxon test. CD4 = CD4+ T-cells; Mono = monocytes; cDC = conventional/myeloid dendritic cells; B = B-cells. **f**, (left) Boxplot showing the classical monocyte D1 response (D1 – D0) in the antigen presentation related gene sets module score (see Methods) using the CITE-seq pseudobulk expression from the same subjects in (**e**). Significance determined by two-tail Wilcoxon test. (right) Bubble plot visualization of the averaged expression of individual leading-edge genes (LEGs) from the antigen presentation related gene set (see Methods) in classical monocytes. **g**, Influenza-specific plasmablast (PB; All HA+ CD27+CD38+CD20^low^CD21^low^; see Methods and Supplementary Information Fig. 3) frequencies at D7 and D0 plotted separately for COVR-F (n = 14), HC-F (n = 15), COVR-M (n = 11), and HC-M (n = 9). Each dot is an individual and the lines connect the D0 and D7 PB frequencies for that individual. Significance of difference between groups is determined by two-tailed Wilcoxon test. **h,** Analysis of the day 28/D0 microneutralization titer fold-change (FC) for each of the four strains in the seasonal influenza vaccine (columns) in female subjects (COVR-F and HC-F) under the age of 65. Each dot represents one individual. The orange and grey lines indicate the average fold change for the HC-F and COVR-F, respectively. P values are derived from generalized regression models accounting for age, race, influenza vaccination history and baseline influenza titers (see Methods). **i**, Similar to (**h**), but for COVR-M and HC-M.

Blood transcriptomic, peripheral blood immune cell frequency, CITE-seq, influenza-specific B-cell, and antibody titer analyses [assessing responses on D1, D7 and day 28 (D28) relative to day 0 (D0)] together pointed to coherent sex-specific innate and adaptive response differences to the vaccine, with COVR-M generally mounting a more potent response than their healthy counterparts and COVR-F (Fig. 2b-2i and Extended Data Fig. 3a-g; Extended Data Tables 8 and 9). These include stronger innate/inflammatory, and in particular, IFN-related transcriptional responses (Fig. 2b, Extended data Fig. 3a) with corresponding greater increases in circulating IFNγ protein levels in serum by D1 in COVR-M (Fig. 2c). Consistent with the hypothesis that this systemic increase in IFNγ may impact diverse cell types expressing the IFNγ receptor and downstream signaling components, single cell CITE-seq analysis revealed that most peripheral immune cells had higher IFN response transcriptional signatures on D1 after vaccination in COVR-M than the other groups (based on comparing D1 vs. D0; Fig. 2d). CD4+ T-cells, B-cells, monocytes and cDCs are shown as examples, all demonstrating stronger IFN responses in COVR-M compared to both COVR-F and HC-M (Fig. 2e). Baseline, pre-vaccination IFN-related transcriptional activity was largely indistinguishable between COVR and HC (Extended Data Fig. 3b). In addition to IFN-related genes, a more robust response was also observed for genes related to antigen presentation including both MHC class I and II genes in classical monocytes of COVR-M (Fig. 2f), likely because IFNγ signaling can increase the transcription of antigen presentation genes^30^. Together, these data indicate that COVR-M mount a stronger circulating IFNγ and corresponding transcriptional response in both innate and adaptive immune cells by D1 following influenza vaccination.

Based on previous studies of influenza vaccination in healthy adults and because heightened innate immune responses elicited by vaccine adjuvants are known to enhance adaptive responses, we hypothesized that the stronger early inflammatory responses in COVR-M would lead to a more robust ensuing humoral response. Indeed, we saw increased D7 B-/plasma-cell related transcriptional signatures in the COVR-M, including when using D7 signature genes reported in independent influenza vaccine studies^29, 31^(Extended Data Fig. 3a,c). Furthermore, COVR-M had a higher increase of influenza-specific plasmablasts than HC-M at D7 (Fig. 2g, Supplementary Information Fig. 3). Consistent with the hypothesis that the stronger early IFN response in COVR-M could potentially help induce a heightened B-cell response, we detected a positive correlation between those two parameters including the extent of influenza-specific plasmablast increases (Extended Data Fig. 3d), which is consistent with previous observations in healthy adults^31^. In agreement with higher influenza-specific plasmablasts in COVR-M, COVR-M have significantly higher influenza-specific antibody responses than HC-M across all but one of the vaccine strains at D28 relative to baseline (Fig. 2h,i; Extended Data Fig. 3e,f; Extended Data Table 9; see Methods). Similarly, the maximum D28 titer across the four vaccine strains was higher in COVR-M than HC-M (Extended Data Fig. 3g), although antibody avidity (as measured by surface plasmon resonance^32, 33^ from D7 and D28 samples) was comparable between COVR and HC in both sexes (Extended Data Table 9). While influenza infection and vaccination history can influence influenza vaccine responses^34^, they alone are unlikely to explain the above findings as the COVR and HC groups had similar baseline antibody titers (Extended Data Fig. 3e,f) and were age/sex-matched and drawn from the same geographic region with very low influenza infection/transmission during the 2020-21 season^35^. Additionally, the statistical model used to assess titer response differences (similar to a model used by the FDA for evaluating influenza vaccine trials; see Methods) incorporated pre-vaccination influenza titers as a covariate (see Methods). The extent of time-dependent immune resolution following COVID-19 was unlikely a factor because TSD and D28 titer responses are not correlated in either sex (data not shown). Together, these observations demonstrate that prior mild infection by SARS-CoV-2 can result in sex-dependent, coordinated changes in both innate and adaptive responses to immunization with non-SARS-CoV-2 antigens months after acute COVID-19.

### GPR56+ CD8+ effector memory T-cells are a source of increased IFNγ in COVID-19-recovered males

Having established that prior mild COVID-19 is associated with both new baseline immune states prior to influenza vaccination (Fig. 1 and Extended Data Fig. 1) and COVR-M-specific responses following vaccination (Fig. 2 and Extended Data Fig. 3), we next attempted to link the two and asked what baseline variables may be contributing to the heightened responses in COVR-M. We focused our analysis on linking the baseline to the increase in IFN-related responses on D1 in the COVR-M because that was the most prominent difference we detected above that could also potentially contribute to the more robust humoral responses in COVR-M (Fig. 3a). Considering that IFNγ is often produced and secreted by T-or NK cells, we first assessed the correlation between the frequency of T- and NK cell subsets from flow cytometry (Supplementary Information Figure 1) and the increase in IFN-related signatures on D1. This analysis suggested that CD8+ T-cells with an effector memory (CD8 EM) phenotype could be a source of IFNγ because the pre-vaccination (D0) frequency of effector-like (CD45RA-CCR7-CD28+ CD27-) CD8+ T-cells correlated with both the increase in circulating serum IFNγ protein and the blood IFN transcriptional signature score in COVR-M between D0 and D1 (Extended Data Fig. 4a, b). We next focused on all the CD8+ T-cells from clusters with an EM phenotype (CD8 EM) in the CITE-seq data based on both surface protein markers and mRNA expression (see Methods and Extended Data Table 10 for the top cluster protein markers). We searched for differences in average surface marker expression of cells in these CD8 EM clusters across the four subject groups and found that GPR56 was the top differentially expressed marker with increased expression in COVR-M relative to the HC-M and COVR-F (Fig. 3b,c; Extended Data Fig. 4c; Extended Data Table 10). This was intriguing because CD4+ EM and TEMRA (EM cells re-expressing CD45RA) T-cells marked by surface GPR56 expression at baseline (before stimulation) have been reported to produce increased amounts of IFNγ upon PMA/ionomycin (PMAI) stimulation^36^. Consistent with this, GPR56+ CD8 EM cells in our data are enriched for a transcriptional signature (derived in an independent study^37^) that marks CD8 EM cells poised to secrete higher levels of IFNγ upon PMAI stimulation (Fig. 3d). Thus, GPR56+ CD8 EM cells could be a potential source of elevated IFNγ production in COVR-M following influenza vaccination. Indeed, the frequency of these cells was elevated in COVR-M relative to both HC-M and COVR-F prior to vaccination (Fig. 3e), but not correlated with the TSD and thus was temporally stable (Extended Data Fig. 4d). Additionally, IFNG transcripts increased significantly in these cells on D1 following influenza vaccination in COVR-M (Fig. 3f, Extended Data Fig. 4e). These data suggest that prior COVID-19 increases the frequency of GPR56+ CD8 EM cells in males and these cells are more poised to make more IFNγ early after influenza vaccination, which together contributed to the higher IFNγ production in COVR-M (Extended Data Fig. 4f); consistent with this hypothesis, this was not observed in GPR56-cells.

**Figure 3.**
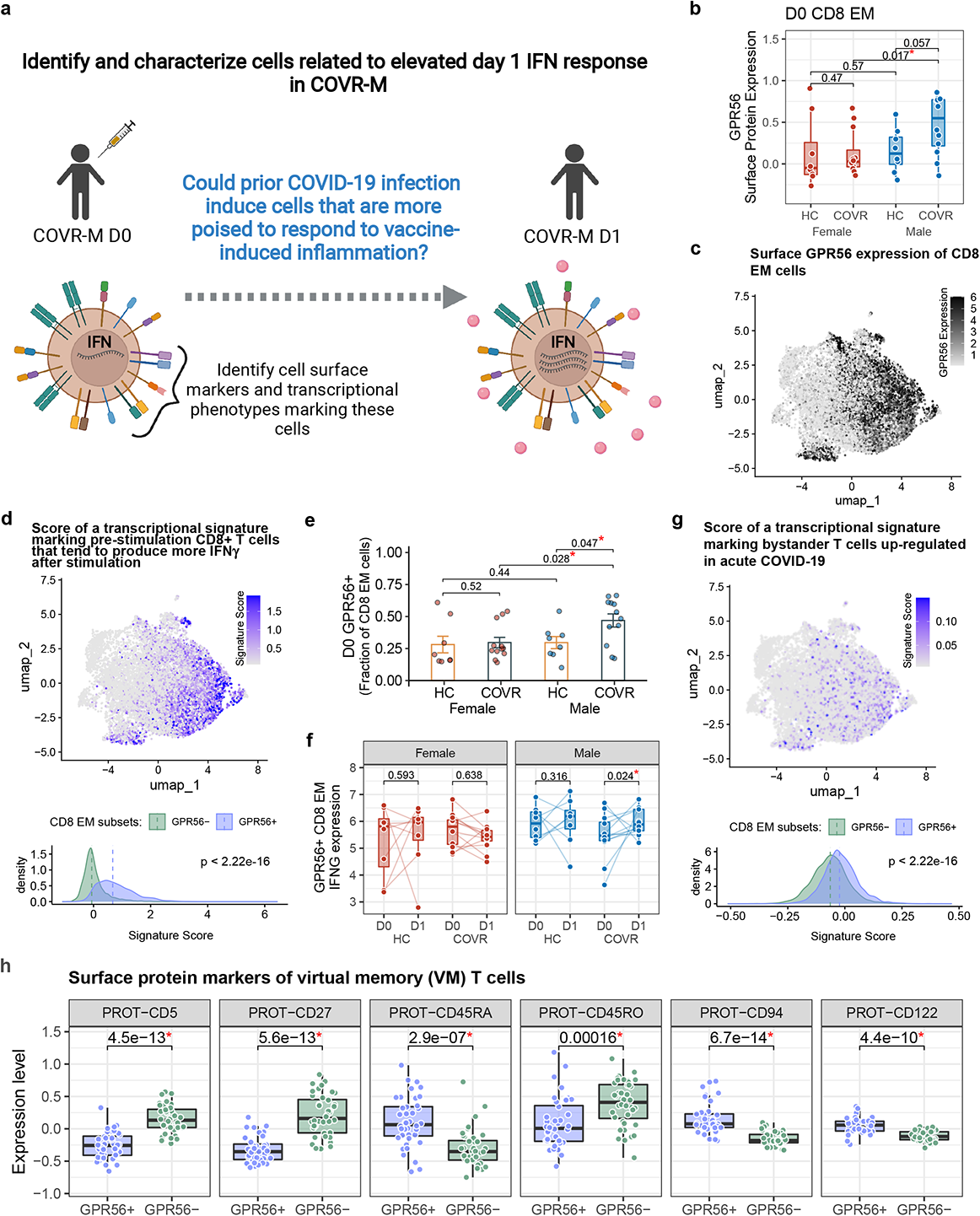
GPR56+ CD8+ effector memory T cells contribute to increased day 1 IFNγ responses in COVID-19-recovered males. **a**, Schematic showing the study questions regarding why COVID-19-recovered males (COVR-M) had elevated early interferon gamma (IFNγ) responses. D0 = day 0, D1 = day 1. **b**, Box plots comparing the sample means of GPR56 surface expression in CD8+ effector memory T-cells (CD8 EM; y-axis) at D0 for COVR females (COVR-F; n = 36), healthy control females (HC-F; n = 24), COVR-M (n = 36), and HC males (HC-M; n = 24). Significance of differences is determined by two-tailed Wilcoxon test. **c**, UMAP visualization of the D0 surface GPR56 protein expression on single CD8+ EM T cells from all 40 subjects with CITE-seq data. UMAP was derived using the top 60 variable surface protein features within the CD8 EM cells (see Methods). **d**, Same UMAP as (**c**) (top) but showing the D0 gene expression signature score computed using genes associated with CD29^hi^ CD8+ T-cells identified earlier in an independent study (Nicolet *et al*^37^, see Methods). This signature at baseline (before stimulation) was found to be associated with increased IFNγ secretion following PMA/ionomycin stimulation. (bottom) Density plot showing the distribution of signature score above in the GPR56+ and GPR56- CD8+ EM T-cell subsets. Dashed line indicates the median of the distribution. Significance of the difference between the medians is determined by two-tailed Wilcoxon test at single-cell level. **e**, Bar plots comparing the proportion of GRP56+ cells (as fractions of CD8+ EM in the CITE-seq single cell data; y-axis) between COVR-F (n = 12), HC-F (n = 8), COVR-M (n = 12), and HC-M (n = 8) at D0. Significance is determined by two-tailed Wilcoxon test. **f**, Box plots comparing D0 and D1 pseudobulk IFNγ gene (IFNG) expression (y-axis) in the GPR56+ CD8+ EM population for COVR-F (n = 12), HC-F (n = 8), COVR-M (n = 12), and HC-M (n = 8). Significance is determined by a linear model accounting for age, race, and influenza vaccination history (see Methods). **g**, Similar to (**d**) but showing the bystander T-cell signature score at baseline (D0) (signature genes originated from Bangs *et al*^39^ and Bergamaschi *et al*^38^, see Methods). **h**, Box plots comparing the average surface expression (y-axis) of the indicated cell surface protein markers for the GPR56+ versus GPR56- CD8+ EM single cells at D0 (n = 40 subjects). Significance is determined by two-tailed Wilcoxon test. Each point represents a subject. Additional markers are shown in Extended Data Fig. 4h.

Mild, non-hospitalized COVID-19 has been reported to induce “bystander activation” (non-SARS-CoV-2 specific) of CD8+ T-cells^38^. Interestingly, the GPR56+ cells are also enriched for a transcriptional signature associated with bystander T-cell activation^38, 39^ (Fig. 3g). In addition, GPR56+ CD8 EM cell frequency is positively correlated with the T-cell activation signature score, which was elevated at baseline in COVR-M as shown above (Fig. 1i, Extended Data Fig. 4g). This suggests that some of these cells may have expanded in a bystander manner during the acute phase of the infection. This prompted us to consider whether these GPR56+ cells are similar to bystander-activated virtual memory (VM) CD8+ T-cells^40^, a feature of which is their ability to be activated rapidly by inflammatory cytokines alone (e.g., IL-12 and IL-18) to produce IFNγ without T-cell receptor (TCR) stimulation^40^. VM CD8+ T-cells develop and expand in the periphery via cytokine stimulation, including IL-15 induced by viral infection (IL-15 concentrations are known to be elevated in acute COVID-19 patients and correlate with disease severity^41, 42^), and subsequently develop a differentiated EM phenotype expressing CD45RA^40^. We assessed several reported surface markers of these cells^40^ in GPR56+ vs. GPR56-cells and found that the GPR56+ cells were indeed phenotypically similar to VM cells (Fig. 3h). For example, GPR56+ cells have higher CD122 but lower CD5 surface expression than their GPR56-counterparts; both markers have been linked to the extent of prior IL-15 (or potentially other inflammatory cytokine) encounters^40, 43^. Interestingly, based on the surface level of CD45RA and CD45RO, the GPR56+ cells appear to situate phenotypically between GPR56- and TEMRA cells (Extended Data Fig. 4h).

Since VM T-cells can be rapidly activated and produce cytokines without clonal, antigen-specific expansion^40^, we assessed the clonality of the GPR56+ CD8 EM cells at different timepoints after influenza vaccination using V(D)J/TCR profiling from the CITE-seq data (combining COVR and HC data in order to maximize the number of available cells). The clonality of both the GPR56+ CD8 EM and TEMRA cells remained stable across days 0 (before vaccination), 1 and 28 following influenza vaccination (Extended Data Fig. 4i,j). The frequencies of GPR56+ CD8 EM clones shared across timepoints within individual subjects were also similar (Extended Data Fig. 4k). Together, these data argue against the notion that the heightened activation of the GPR56+ cells early after influenza vaccination in COVR-M was due solely to TCR-dependent T-cell activation and clonal expansion. As was shown previously^40, 44^, a more plausible explanation is that these CD8+ VM-like cells were activated to produce IFNγ by the inflammatory cytokines elicited by the influenza vaccine (e.g., via IL-12 or IL-6 produced by monocytes or dendritic cells after they were stimulated by components of the influenza vaccine)^45^ in an antigen-independent manner. Despite their resemblance to VM cells, some of the GPR56+ cells could have developed from naïve cells via conventional, non-bystander pathways – e.g., some could be developed during acute COVID-19 and are specific for SARS-CoV-2, although none of these cells had a CDR3 sequence that matches a public clone deemed to be specific for SARS-CoV-2 (data not shown). Bona fide, antigen-specific memory CD8+ T-cells developed from naïve cells via TCR stimulation have also been shown to produce IFNγ in response to inflammatory cytokines alone in mice^46, 47^. In addition, NK cells may also be a contributor: IFNG transcript was increased more in COVR-M than HC-M and COVR-F on day 1 after influenza vaccination in CD16^lo^ NK cells (p<0.05; data not shown).

Taken together, we demonstrate a population of CD8 EM T-cells marked by GPR56 expression with antigen-agnostic pro-inflammatory potential after heterologous vaccination. Importantly, these cells emerged in otherwise clinically healthy individuals and are especially elevated and more poised to respond in males who were months recovered from mild SARS-CoV-2 infection, providing additional evidence for sex-specific, functionally relevant immune set points linked to prior mild COVID-19.

### Partial “reversal” of gene expression imprints in circulating monocytes following influenza vaccination

Given the potential for long-lasting vaccine-induced “training” effects^2, 48, 49^, we next asked whether influenza vaccination may help “reverse” the post-COVID-19 immune states back towards that of healthy controls (pre-vaccination) who never had COVID-19 (“the healthy baseline”, Fig. 4a). We focused on the monocytes because of their depressed transcriptional state (in COVR vs. HC) involving genes such as pattern recognition receptors (Fig. 1f,g) and because vaccines can potentially induce long-lasting changes in these cells^2^. By using the HC baseline (D0) as a healthy reference, we used CITE-seq data to assess the average expression of these depressed genes identified earlier before and after vaccination in COVR subjects, assessing this separately for classical (Fig. 1f) and non-classical monocytes (Fig. 1g) in males and females (Extended Data Fig. 5a,b). As was observed above, these genes had lower average expression in COVR than HC in both sexes at D0 before vaccination. However, their average expression increased towards that of the HCs by D1 and persisted through D28 in both COVR-F and COVR-M, although the effect appeared stronger in COVR-F (Extended Data Fig. 5a,b).

**Figure 4.**
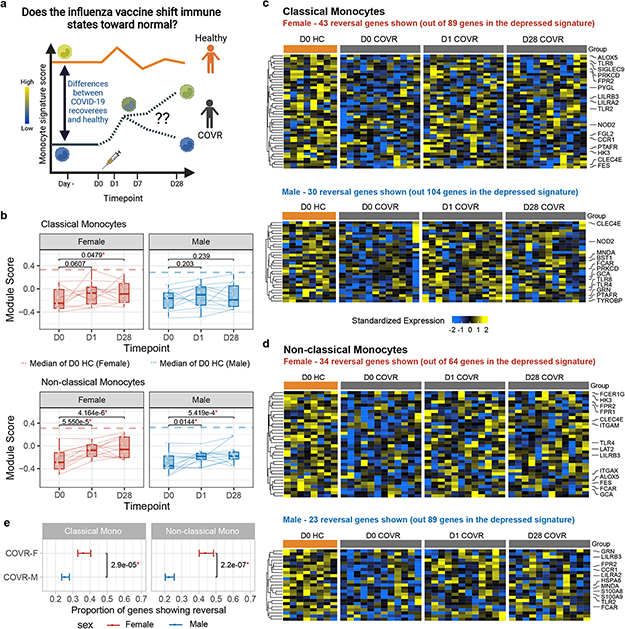
Partial reset of post COVID-19 gene expression imprints in monocytes by influenza vaccination. **a**, Schematic showing the study questions explored. D0 = day 0, D1 = day 1, D7 = day 7, D28 = day 28. **b**, Box plots showing the repressed monocyte signature module scores (y-axis) in COVR-F (n = 12) and COVR-M (n = 12) at D0, D1 and D28 using the CITE-seq pseudobulk gene expression data in classical (top) and non-classical (bottom) monocytes. The gene set comprising the signature includes the union of the BTM-M4.0 and M11.0 leading-edge genes for that cell type (see Fig. 1f,g). See also Extended Data Table 5. The dashed line represents the median D0 score of the HCs of the same sex. Lines connect data points from the same subject at different timepoints. Significance of differences is determined by a mixed-effects model with age and race as fixed effects and subject as random effect. **c**, Heatmap showing the expression of the “reversal” genes in classical monocytes. Reversal genes are defined as those genes in the baseline repressed signature (see Fig. 1f) whose expression in COVR subjects at D1 and D28 after vaccination moved towards the baseline (pre-vaccination) expression of HCs. COVR-F (top) and COVR-M (bottom) shown separately; D0 HC are also included for comparison (see Methods for the definition of reversal). The rows are genes and columns are individual samples (grouped by subject/timepoint combination) with timepoint and subject group labels shown at the top, including HC-F D0 (n = 8), HC-M D0 (n=8), COVR-F (n = 12 at each timepoint), and COVR-M (n = 12 at each timepoint). The names of genes that belong to gene sets of functional interest (Gene Ontology (GO): pattern recognition receptor activity and GO: myeloid leukocyte activation) are shown. Gene expression values are shown as row-standardized z-scores. **d**, Similar to (**c**) but for non-classical monocytes (see Fig. 1g for the repressed signature in non-classical monocytes). **e,** Comparison of the proportion of repressed monocyte and non-classical signature genes (see Fig. 1f,g respectively) that show reversal in in COVR-F versus COVR-M (see Methods for the definition of reversal). The mean and 95% confidence intervals (denoted by the bars) are shown with the latter derived from a bootstrapping procedure (see Methods). Significance is determined by the two-tailed Wilcoxon test between the bootstrapped samples.

Quantifying the average expression (module score) of these sex- and cell type-dependent gene sets as identified earlier (Fig. 1f,g) within individual subjects over time confirmed a similar significant trend of reversal towards the HCs (Fig. 4b). This analysis further revealed that the extent of this partial reversal in gene expression was more pronounced in the non-classical than the classical monocytes (Fig. 4b). We next identified the individual genes within these depressed gene sets that showed significant reversal based on the changes they demonstrated on days 1 and 28 compared to baseline (see Methods). In both classical and non-classical monocytes, the fraction of reverting genes was significantly higher in females than males (Fig. 4c-e), although several TLRs (e.g., TLR2, TLR4) and NOD2 showed significant reversal in both males and females in one or both monocyte subsets (Fig. 4c,d). These changes were unlikely due to natural immune resolution following infection because the baseline (D0) expression of these genes did not correlate with TSD across COVR subjects (Extended Data Fig. 1h), they increased acutely by D1 following vaccination, and they persisted to D28 (and potentially even beyond). Interestingly, unlike our depressed signature (Fig. 1f, g, Extended Data Fig. 1i), other monocyte related transcriptional signatures known to have lower expression during acute COVID-19, such as genes related to antigen presentation, inflammatory and NF-kB activation, and myeloid suppressor cells^19–24^, were largely similar between COVR and HC at D0/baseline; vaccination also did not consistently elicit longer-lasting changes in these signatures out to D28 (Extended Data Fig. 5c,d). Together, CITE-seq analysis revealed that the early (D1) response to influenza vaccination elevates a set of previously (i.e., before vaccination) depressed innate immune receptor/defense genes in the monocytes of COVR subjects out to at least D28 post vaccination. These data suggest that the early inflammatory responses to influenza vaccination helps to revert the post-COVID-19 immune state of monocytes back towards that of healthy, particularly in COVR-F.

## Discussion

While both acute and long-term immune perturbations in hospitalized COVID-19 patients have been reported^24, 50–55^, less is known regarding healthy recovered individuals with prior mild, non-hospitalized SARS-CoV-2 infection months after acute illness. Furthermore, most studies of post-COVID-19 have focused on adaptive and antigen-specific immunity. Here we reveal that prior mild, non-hospitalized COVID-19 in otherwise healthy individuals is associated with sex-specific immune imprints beyond SARS-CoV-2 specific immunity, some of which only become apparent after heterologous challenge via influenza vaccination (i.e., a vaccine that is antigenically distinct from SARS-CoV-2). Thus, COVID-19 has the potential to impact the response to future immunological perturbations long after acute disease and convalescence. This is of public health importance given that the majority of the more than 500 million global SARS-CoV-2 infections have been mild and not required hospitalization^56^. The few studies of convalescent, mild COVID-19 have included patients with multiple medical co-morbidities, relatively small sample sizes, and did not evaluate sex-specific effects^57–59^. To our knowledge, ours is the first study using multimodal single cell and systems immunology approaches to reveal sex-specific molecular and cellular immune imprints and future immune response differences associated with prior mild COVID-19 in otherwise healthy individuals, particularly those without confounding comorbidities such as autoimmunity or immunodeficiency.

Our findings are consistent with the sex dimorphic nature of acute responses to SARS-CoV-2 and other immune challenges^9–14, 16, 17, 60^. Females are generally more susceptible to autoimmunity and tend to mount heightened inflammatory responses to infections and vaccines^61^; it was therefore surprising to find the qualitative opposite here in which COVR-M were found to have a more “activated” immune status at baseline and stronger innate and adaptive responses to influenza vaccination. While some of these might be attributable to differences in acute disease severity (e.g., males tended to have more severe disease than females), it is not clear how that might have manifested in our mild, non-hospitalized patients as neither the self-reported duration of acute illness nor antibody titers against SARS-CoV-2 were different between COVR-M and COVR-F (duration of illness Wilcoxon p=0.37; USA-WA1 IC_50_ Wilcoxon p=0.26), which together suggest that our observations are potentially independent of severity or immune response quality during acute disease. Persistent immune state changes (over months) in patients with “long COVID” have recently been reported^51, 62^, but most of the individuals in our study reported no or minor post-COVID-19 sequelae. Thus, immunological modifications with functional consequences can still be present after clinically resolved, mild COVID-19. Although our study found heterologous vaccine response benefit in COVR-M (e.g., elevated influenza vaccine titers), the impact of prior mild COVID-19 on other perturbations such as non-SARS-CoV-2 respiratory infections remains to be determined. For example, airway neutrophil inflammation before respiratory syncytial virus exposure is associated with symptomatic outcomes^63^. As future work it could also be informative to assess whether some of the sex-specific imprints, including the differences in heterologous vaccination responses identified here, are associated with clinical sequelae present in those with “long COVID”^1^.

The sex-specific post-vaccination cellular and molecular dynamics observed in this study suggest that the more “primed” baseline immune states in COVR-M could have helped establish the more robust IFN, plasmablast, and antibody responses on days 1, 7, and 28, respectively, following influenza vaccination, which is antigenically distinct from SARS-CoV-2. These observations are consistent with earlier findings that the heterologous (non-antigen-specific) effects of vaccination (e.g., BCG) can be sex-specific^64^. However, instead of myeloid cells such as “trained” monocytes, a poised baseline state within COVR-M could be attributed to a subset of GPR56+ CD8 EM T-cells with a surface phenotype that resembled virtual memory cells. Interestingly, a qualitatively similar “priming” effect has also been observed in repeated homologous vaccination, such as increased innate responses following the second dose of the Pfizer-BioNTech COVID-19 vaccine or the AS01-adjuvanted hepatitis B vaccine compared to the first dose^65, 66^. It remains to be seen whether similar, non-antigen specific virtual memory CD8 EM T-cells are involved, particularly given that these homologous (repeated dosing) vaccine-induced responses did not appear to be sex-specific and the second dose was given only 3-4 weeks after the first (compared to the months between mild COVID-19 and influenza vaccination in our study).

Changes in the transcriptional and epigenetic profiles of peripheral monocytes have been described in both acute and convalescent COVID-19 patients with moderate-to-severe disease, but few included patients months out from infection^21, 23, 53, 55, 67^. These previously described changes during acute disease include the depressed inflammation/antigen-presentation transcriptional phenotypes that are distinct from the monocyte depression that we detected months post COVID-19. This monocyte imprint involving transcriptionally depressed innate defense/receptor genes is consistent with the notion of trained immunity^2^. However, our signature likely reflects different biology than the “poised” trained monocytes (based on epigenetic and *in vitro* stimulation studies) found in an earlier study of seven COVID-19-recovered patients, probably because those were hospitalized patients with more severe acute disease (e.g., most had pneumonia) and the time since discharge was relatively short (∼4-12 weeks)^55^. Our signature was also distinct from the other depressed antigen presentation or myeloid suppressor cell like states found in acute COVID-19. The finding that the monocyte depression imprint we detected was partially reversible by seasonal influenza vaccination suggests that in addition to providing antigen-specific protection, vaccines could help reset certain immune cell states in an antigen-agnostic manner. It is possible that the distinct biological mechanisms that facilitate different aspects of trained innate immunity (e.g., innate immune cells can exhibit both enhanced or repressed responses to antigenic challenges after a preceding antigenic or inflammatory exposure^2, 68^) may also drive both the persistent post-COVID-19 monocyte repression and subsequent lessening of that repression following influenza vaccination. Given that females tend to mount stronger innate/inflammatory responses to vaccination than males, similar mechanisms might have driven the stronger vaccine-induced reversal detected in COVID-19-recovered females^61^. As trained innate immunity can be mediated through epigenetic and metabolic changes within cells, future studies could explore these potential mechanisms using this and other cohorts, including the influences of sex/gender, acute disease severity, and age among subjects with a range of post-COVID clinical sequelae.

T-cell bystander activation has been reported following natural viral infections^69^, including SARS-CoV-2^38^. More recently, bystander activated CD8 EM T-cells have been identified as playing an important role in controlling early infection, including VM cells that have no prior antigen exposure or TCR engagement^40, 44^. As these cells can emerge following cytokine stimulation alone, it is possible that a stronger or more prolonged cytokine response to SARS-CoV-2 in males relative to females may have resulted in the elevated frequencies of both the GPR56+ CD8+ VM-like cells in COVR-M^70^. This hypothesis is consistent with reports that males hospitalized with COVID-19 tend to experience greater innate immune activation (as measured by circulating cytokines) compared to females^60, 71^.

Some of the immune state imprints we observed could be shared among different types of viral infections (e.g., given the involvement of similar inflammatory cytokines like IL-15 in the acute response), but some are likely unique to SARS-CoV-2, as shown in our preliminary comparison with natural influenza infection. Features such as the route of the infection and viral tropism are potentially important determinants, although a detailed comparison with influenza or other viral infections is outside the scope of the current work. Our findings point to the possibility that any infection or immune challenge may change the immune status to establish new set points. Thus, the immune status of an individual is likely shaped by a multitude of prior exposures and perturbations. Our study’s unique design contributes data and conceptual advances to help reveal underlying principles regarding what happens after two well-defined natural immunological encounters in humans: mild COVID-19 and influenza vaccination. Thus, our observations provide a basis for the development of hypotheses regarding more complex scenarios including what happens over longer timescales with additional encounters.

**Limitations of this study** include most study subjects were younger than 65 and thus these findings may not apply to the elderly, an important population of COVID-19 recoverees. Additionally, our findings are largely associative in nature and the study design does not allow the linking of acute response phenotypes to the long-term imprints in the same individuals. Some of the imprints we considered as stable given lack of association with TSD may still be evolving slowly (or could be limited by statistical power for detecting association with TSD). And while there was no clear difference in disease severity or duration between the COVID-19-recovered males and females in our study (and no subjects were hospitalized), it is possible that our sex-specific findings reflect unappreciated clinical factors. It is possible that some of the post-vaccination reversal towards the healthy, pre-vaccination state by D28 may also in part be due to ongoing disease resolution. However, this is unlikely the case for the vaccine-induced elevation in the expression of the reversal genes towards the healthy state because those changes were clearly detectable on D1 after vaccination and persisted through D28, especially in females, indicating that this reversal was driven (or at least accelerated) by vaccination and could not be attributed to the “natural” resolution process alone. At the time of the study, the COVID-19 vaccine was not yet available and circulating viruses were limited due social distancing and face mask adherence in the local area^72^; this makes us more confident that the observed immunological changes were due to COVID-19 and not attributable to other vaccines or viral infections between acute COVID-19 and influenza vaccination.

While individual exposure and immune “training” history beyond prior COVID-19 likely play an important role in shaping the responses to influenza vaccination at the individual level, our study design assessed imprints at the group level. We enrolled COVID-19-recovered subjects together with matching HCs who never had COVID-19 at the time of influenza vaccination. It is reasonable to believe that the extent of heterogeneity in the exposure history within each group is, on average, quite comparable between the COVR and matched HC groups. Thus, the group level features shared across individuals (e.g., depressed monocyte signatures or the heightened IFN response after influenza vaccination in COVR-M) that we were able to detect should be largely independent of differences in personal exposure history. This approach thus allowed us to robustly uncover both pre-vaccination (baseline) and immune response features associated with prior mild COVID-19.

Logistical and pandemic-related challenges prevented us from enrolling and longitudinally following individuals from acute COVID-19 through influenza vaccination. As future work it could be informative to link acute immune responses to the long-term imprints we revealed in this study. While it would be informative to further assess our findings in follow up cohorts, given our observation that vaccination could perturb some of the immune imprints associated with prior mild COVID-19, identification and recruitment of a sufficient number of individuals who have not had a viral infection or COVID-19 vaccines since their COVID-19 disease would be impractical. Our study is thus unique in allowing us to dissect the immune imprints in healthy recoverees following the first wave of COVID-19 without apparent interference from perturbations between acute disease and influenza vaccination, including COVID-19 and other vaccines. Finally, the functional and clinical implications of the vaccine-induced partial reversal of the depressed gene signatures in monocytes remain to be determined.

The influenza vaccine was selected for this study due to its public health importance and the well understood immune response dynamics in blood that permitted sample collection at the most informative time points post-vaccination. Subjects in this study had previous exposure to influenza vaccination and/or natural infection that complicates interpretation of antibody responses. However, we were able to account for existing influenza antibody titers in the analyses. Administration of a vaccine antigen to which study participants were naïve (e.g., rabies or yellow fever vaccine) would have removed potential confounding due to the aforementioned prior influenza exposures, but the response dynamics to these vaccines are less well characterized and there was a weak public health indication to administer these vaccines compared to influenza vaccination. Tissue-level or lymph node responses were not assessed in our study due to logistical and clinical challenges, especially during the first wave of the pandemic in 2020, but can provide additional important insights.

Despite these limitations, our work provides conceptual advances regarding how even mild viral infections can stably shape human immune statuses and functions long-term after acute illness, thus establishing new antigen agnostic baseline set point with potential impacts on future responses^73^, and in turn, how heterologous vaccination can reveal such imprints and potentially help reverse the immune system back towards the state before SARS-CoV-2 infection.

### Data Availability

Raw and processed data from the whole blood bulk RNA-seq and single-cell CITE-seq are available from the NCBI Gene Expression Omnibus, accession numbers GEO: GSEXXXXXX (https://www.ncbi.nlm.nih.gov/geo/query/acc.cgi?acc=GSEXXXXXX) and GEO:GSEXXXXXX (https://www.ncbi.nlm.nih.gov/geo/query/acc.cgi?acc=GSEXXXXXX), respectively (will be released to public at time of publication). Additional datasets, including clinical, proteomics, flow cytometry, and influenza antibody measurements, are available at: https://doi.org/10.5281/zenodo.5935845 (will be released to public at time of publication). The influenza infection dataset we utilized was downloaded directly from GEO: GSE68310 (https://www.ncbi.nlm.nih.gov/geo/query/acc.cgi?acc=GSE68310).

### Code Availability

Analysis code, extended patient and sample metadata are available at: https://github.com/niaid/covid-flu (will be released to public at time of publication).

## Data Availability

Raw and processed data from the whole blood bulk RNA-seq and single-cell CITE-seq will be made available to the NCBI Gene Expression Omnibus at the time of journal publication. Additional datasets, including clinical, proteomics, flow cytometry, and influenza antibody measurements will also be released to the public at time of publication. The influenza infection dataset we utilized was downloaded directly from GEO: GSE68310 (https://www.ncbi.nlm.nih.gov/geo/query/acc.cgi?acc=GSE68310).

## Acknowledgement

We thank the study subjects for their participation. We thank Kaitlyn Sadtler for assistance with the development of the Cytek 36-color flow cytometry panel. We thank Ronald Germain for critical review of the manuscript. Fig. 1a, 1b, 2a, 3a, 4a, Extended Data Fig. 2a were created using BioRender.com. This research was supported by the Intramural Research Programs of the NIAID and the Intramural Programs of the NIH Institutes supporting the NIH Center for Human Immunology. The content of this publication does not necessarily reflect the views or policies of the Department of Health and Human Services, nor does mention of trade names, commercial products, or organizations imply endorsement by the U.S. Government.

## Consortia authors

OP11 Clinical Staff: Princess Barber, Valerie Mohammed, Cindy Palmer, Anne Carmona, Jean Hammer, Deitra Shipman

---

**Extended Data Figure 1.**
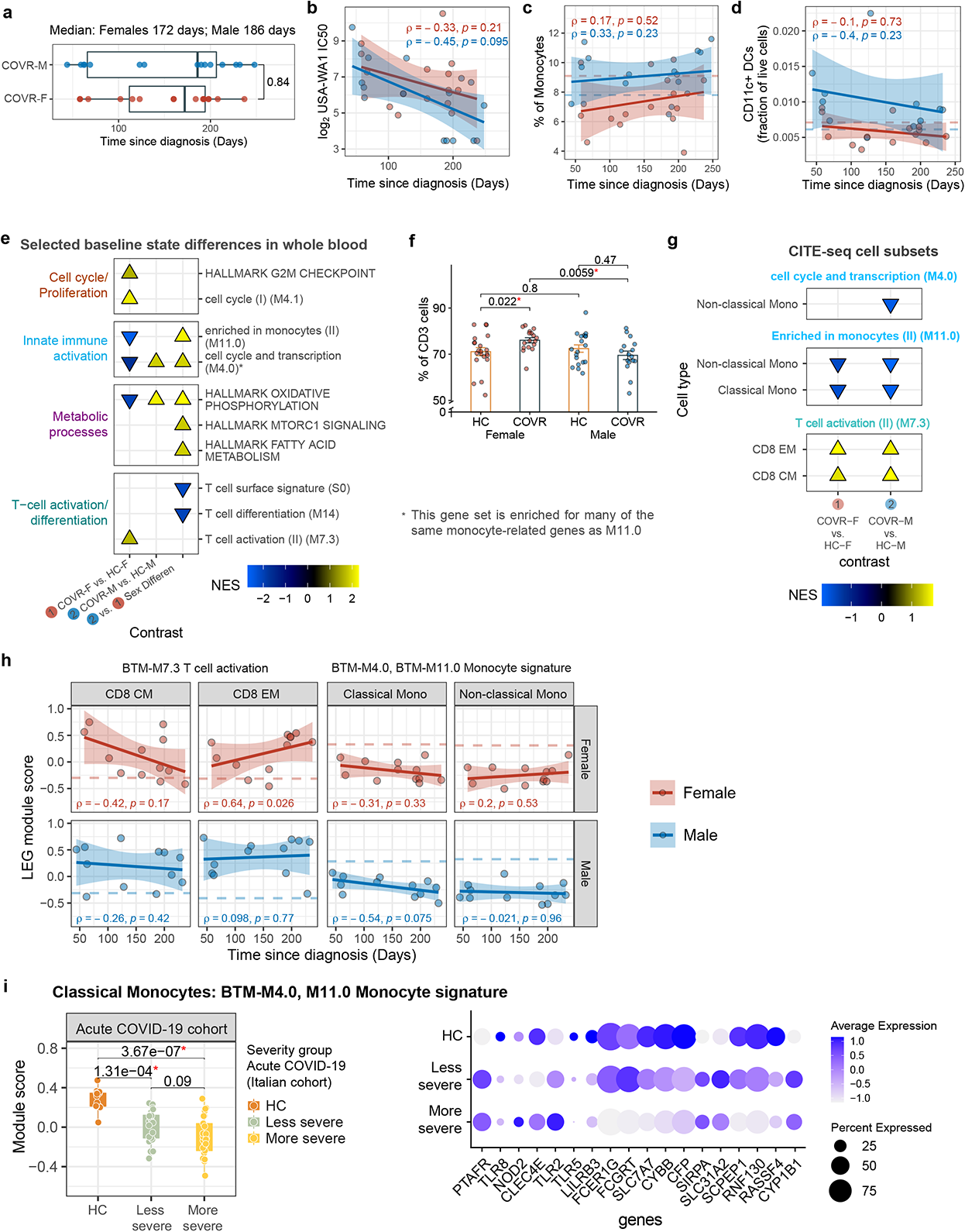
Baseline (pre-vaccination) molecular and cellular differences between COVID-19-recovered subjects and healthy controls. **a**, Box plot showing the distribution of time since diagnosis (TSD; x-axis) in COVID-19-recovered (COVR) females (COVR-F; n=16) and COVR males (COVR-M; n=15). Two participants with asymptomatic COVID-19 infection and thus unknown TSD are not included. Significance of group difference is determined by two-tailed Wilcoxon test. **b**, Scatterplot showing the correlation between the TSD (x-axis) and the SARS-CoV-2 neutralization titer (WA1 strain; y-axis) for COVR subjects at day 0 (D0) prior to influenza vaccination. Spearman’s rank correlation and p values are shown. Participants with asymptomatic COVID-19 infection not included in TSD analyses. **c**, Similar to (**b**), but for the percentage of monocytes in peripheral blood as measured by the complete blood count (y-axis) at D0. **d**, Similar to (**b**), but for the proportion of CD11c+ dendritic cells (DCs; as fraction of live cells from flow cytometry; y-axis) as measured by flow cytometry of PBMCs at D0. **e**, Blood transcriptomic analysis of the stable baseline (before influenza vaccination) differences among COVR and HC groups. Enrichment plot shows the normalized enrichment scores (NES) of selected gene sets of the different comparisons (GSEA FDR < 0.05; see Methods; see Extended Data Table 4 for all significant gene sets with FDR < 0.05). The NES are plotted separately for COVR-F versus HC females (HC-F), COVR-M versus HC males (HC-M), or the difference between the two sets of comparisons (COVR-M versus COVR-F taking healthy sex differences into account). Positive (negative) NES indicates that gene set scores are higher (lower) in the first group than the second group listed in the comparison. Only gene sets not correlated with TSD across COVR subjects at baseline are considered stable. **f**, Bar plots comparing the percentage of CD3+ cells (T-cells) between COVR-F (n = 17), COVR-M (n = 16), HC-F (n = 21), and HC-M (n = 19) at baseline (average of day -7 and D0) as measured by whole blood clinical flow cytometry data covering T-and B-lymphocytes and Natural Killer cells (TBNK; y-axis). Significance of pairwise differences shown is determined by two-tailed Wilcoxon test. **g**, Similar to (**e**), but for a subset of monocyte and T-cell activation gene sets with significant enrichment (p < 0.05) using the D0 CITE-seq pseudobulk expression for the specified cell types (see Methods; see Extended Data Table 6 for complete results). **h**, Scatterplots showing the relationship between the TSD and leading-edge gene (LEG) module scores [left two boxes: the T-cell activation gene set (BTM-M7.3); right two boxes: the union of the LEGs from monocyte gene sets BTM M4.0 and M11.0; see Methods] in COVR-F (n = 12) (top row) and COVR-M (n = 12) (bottom row) at D0 using the CITE-seq pseudobulk data of the indicated cell types. Each dot represents a COVR subject. The dotted lines represent the median score for the sex-matched HC group at D0 in the comparison shown. Spearman’s rank correlation and p values are shown. **i**, (left) Box plot comparing the classical monocyte pseudobulk module scores of the LEGs used in Fig. 1f (union of female (F) and male (M) gene sets) in an acute COVID-19 CITE-seq dataset from Liu *et al*^19^. Both M (n = 50) and F (n = 9) subjects are included in all three groups (HC n = 13, less severe n = 21, more severe n = 25). Each dot represents a sample. P values from the indicated two-group comparisons are shown. P values were generated using the moderated T statistics from linear model of the limma package in which samples from the same donors were treated as duplicates (See Methods). (right) Bubble plot showing expression of the genes in Fig. 1f right panel within the classical monocyte CITE-seq data from Liu *et al* in the same three patient groups shown in the left panel.

**Extended Data Figure 2.**
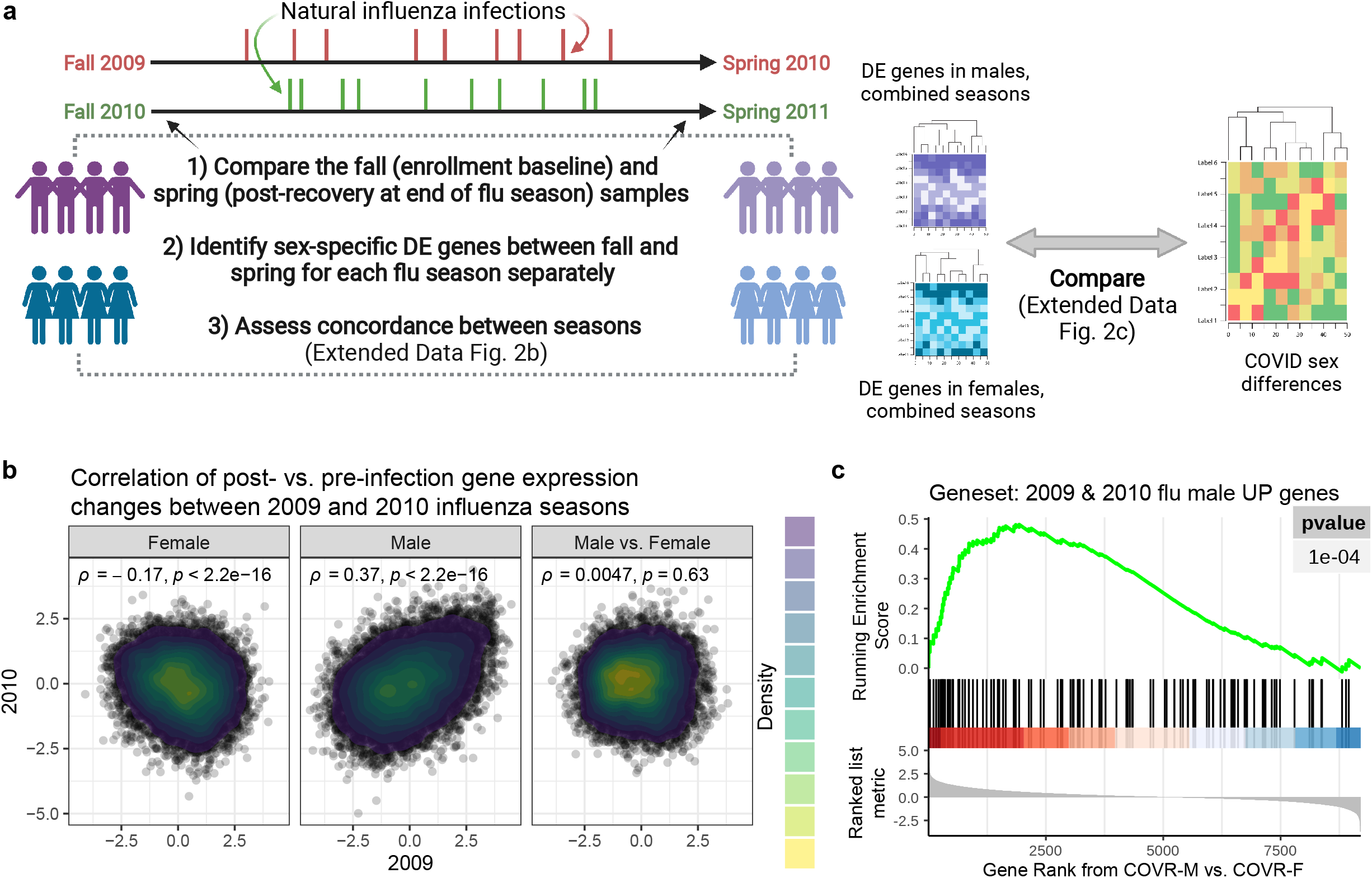
Persistent post-infection gene expression changes following natural influenza infection. **a**, Schematic showing the approach used to evaluate changes in blood gene expression between before (pre-infection baseline) and months after natural influenza infection over two distinct seasons published in Zhai *et al*^25^, and how those gene changes may relate to sex-specific differences resulted from prior COVID-19 in this study. **b**, Scatter density plot showing the correlation between the gene expression changes (see Extended Data Table 7) before (fall) and after (spring) natural influenza A infection in 2009 (x-axis) and 2010 (y-axis) for females (F; left), males (M; center), and M vs F contrast (right). Shown are Spearman’s rank correlation and p values. **c**, Gene set enrichment plot of the genes that are upregulated in M between fall (pre-infection) and spring (post-infection) in both 2009 – 2010 and 2010 – 2011 seasons. Genes were ranked by the signed log10(p-value) in the COVID-19-recovered (COVR)-M vs COVR-F contrast at baseline using only subjects under 65 years of age. The tick marks denote the location of the genes in the influenza gene set.

**Extended Data Figure 3.**
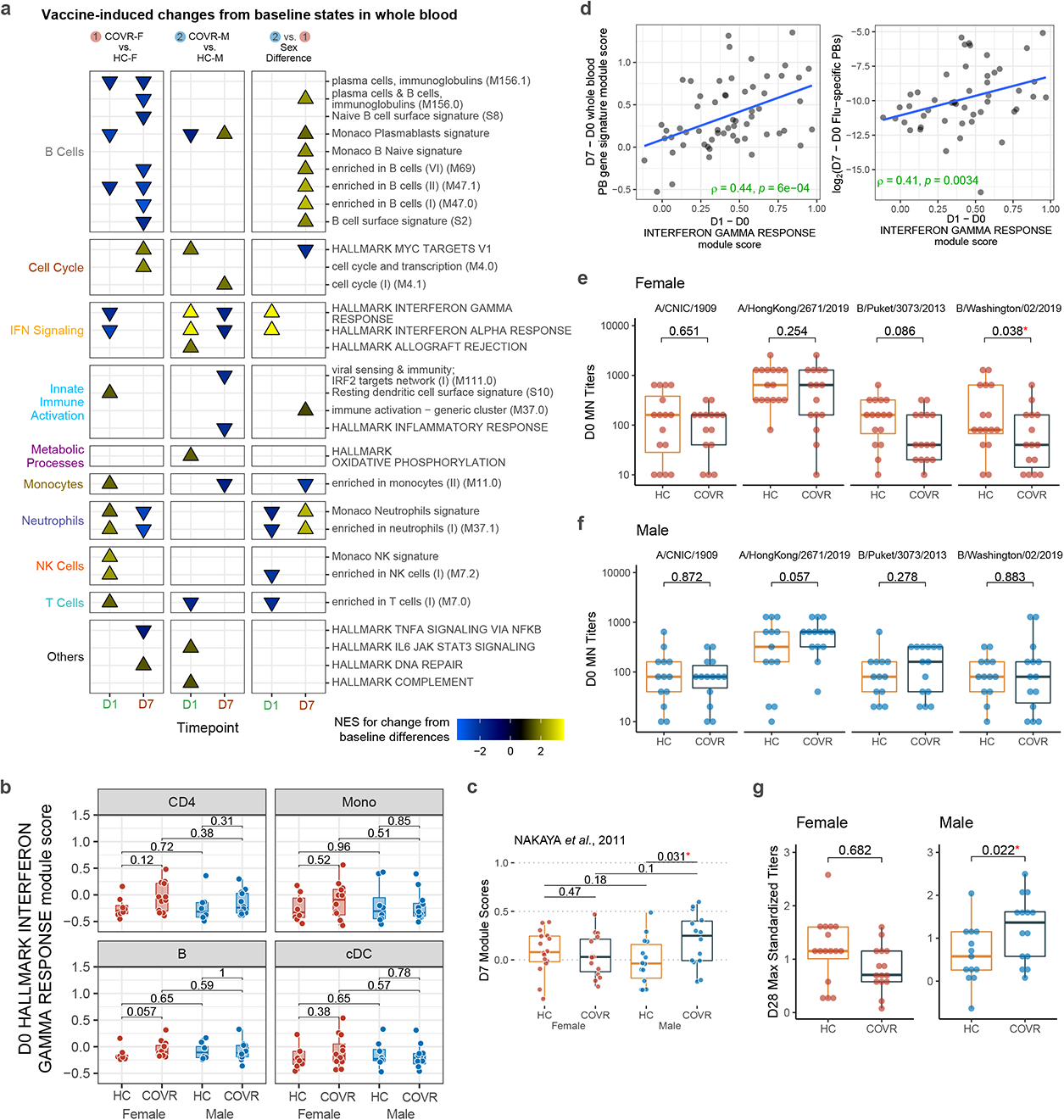
Sex-specific molecular, cellular, and humoral response differences to influenza vaccination between COVID-19-recovered individuals and matching controls. **a**, Similar to Extended Data Fig. 1e but here showing enriched gene sets in whole blood comparing the early [day 1 (D1) and day 7 (D7)] influenza vaccination responses in COVID recovered (COVR) vs. healthy control (HC) subjects for females (F; Contrast 1), males (M; Contrast 2), and sex differences (Contrast 2 vs. 1; i.e., COVR-M versus COVR-F taking healthy sex differences into account; see Methods). Plotted are the gene sets that show significant changes from the baseline [day -7 and day 0 (D0)] within each comparison group [e.g., COVR-F and HC-F for D1] and significant differences between the two groups at the indicated timepoints (FDR < 0.05; see Extended Data Table 6). **b**, Similar to Fig. 2e, but showing the D0 Hallmark Interferon Gamma Response module score for the indicated cell types from the CITE-seq pseudobulk expression data. CD4 = CD4+ T-cells; Mono = monocytes; cDC = conventional/myeloid dendritic cells; B = B-cells. **c**, Box plot showing the D7 whole blood signature score from genes identified in Nakaya *et al*^31^ whose D7/D0 fold-change positively correlated with day 28 (D28) influenza hemagglutination inhibition titers. Only subjects under 65 years of age [COVR-F (n=15), COVR-M (n=14), and HC-F (n=16), and HC-M (n=14)] are included. Significance of differences is determined by two-tailed Wilcoxon test. **d**, Scatterplot showing the correlation of the whole blood D1 – D0 Hallmark Interferon Gamma Response gene set module score (x-axis) to the whole blood D7 – D0 plasmablast (PB) gene set module score (left y-axis; Monaco *et al*^74^) and D7 – D0 difference of influenza-specific PB (all HA+ CD27+CD38+CD20^low^CD21^low^) frequency from flow cytometry (right y-axis; as fraction of CD19+ B-cells). Only study participants < 65 years of age are included. Spearman’s rank correlation and p values are shown. **e**, Box plots showing the D0 (pre-vaccination) microneutralization titers (y-axis) for each of the four strains in the seasonal influenza vaccine (columns) in females (COVR-F and HC-F) under the age of 65. P values are from linear models accounting for age and race (see Methods). **f**, Similar to (**e**) but for males (COVR-M and HC-M) under 65 years of age. **g**, Maximum standardized influenza vaccine titer (among the four strains in the vaccine; y-axis) at D28 after vaccination for females (left) and males (right), respectively. Statistical significance of COVR vs. HC difference was determined by linear regression models accounting for age, race, influenza vaccination history, and baseline influenza titer (see Methods).

**Extended Data Figure 4.**
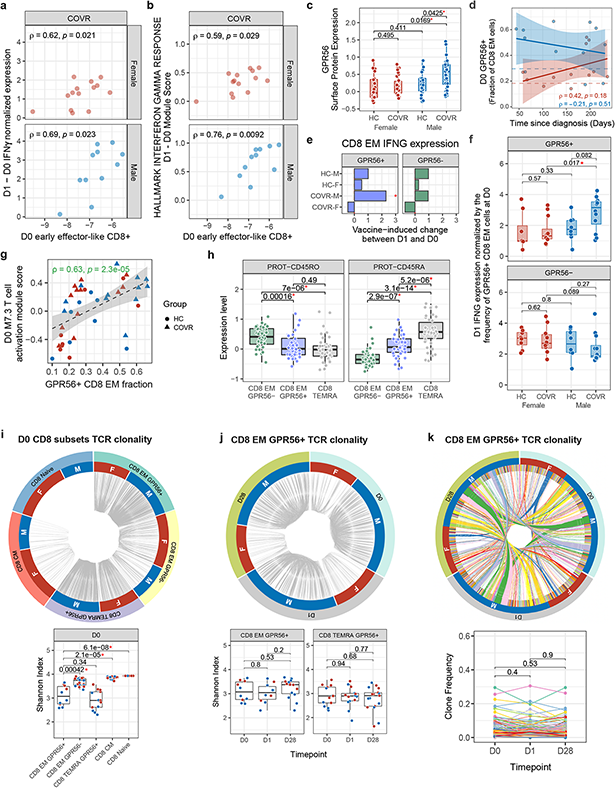
GPR56+ virtual memory-like CD8+ T-cells contributes to increased day 1 IFNγ response in COVID-19-recovered males. **a**, Scatterplots showing the correlation between the day 0 (D0) log_2_ frequency of early effector-like CD8+ T-cells measured by flow cytometry (as fractions of live lymphocytes; see Population 50 in Supplementary Information Table 1 and Supplementary Information Fig. 1; x-axis) and the change (D1 – D0) in serum interferon gamma (IFNγ) protein levels measured by the OLINK platform (y-axis) for COVID-19-recovered (COVR) females (COVR-F; top, n = 14) and COVR males (COVR-M; bottom; n = 11). Spearman’s rank correlation and p values are shown. **b**, Similar to (**a**) but showing the correlation between the D0 log_2_ frequency of early effector-like CD8+ T-cells measured by from flow cytometry (as fraction of live lymphocytes; x-axis) and the change (D1 - D0) in the whole blood signature score of the Hallmark IFNγ Response gene set (y-axis). **c,** Similar to Fig. 3b but for samples from all available timepoints, including D0, D1, and day 28 (D28). Significance of differences is determined by a mixed-effects model with age and race as fixed effects and subject as random effect. **d**, Scatterplot showing the correlation between the frequency of GRR56+ cells at D0 [as fraction of CD8+ effector memory T-cells (CD8 EM; y-axis)] and the time since diagnosis (x-axis) for COVR subjects in the CITE-seq cohort [COVR-F (n = 12) and COVR-M (n = 12)]. Dashed lines show the median frequency for the healthy control (HC) groups. Spearman’s rank correlation and p values are shown. **e**, Bar plot showing the T statistic of the vaccine-induced change (D1 - D0) in IFNγ gene (IFNG) expression using CITE-seq pseudobulk data (x-axis) within the GPR56+ and GRP56-CD8 EM for HC-F (n= 8), COVR-F (n = 12), HC-M (n = 8), and COVR-M (n = 12). * p < 0.05 from mixed effect model correcting for age, race, and influenza vaccination history (see Methods). **f**, Boxplots showing the D1 normalized IFNG mRNA expression in GPR56+ cells (y-axis) in HC-F (n = 8), COVR-F (n = 12), HC-M (n = 8) and COVR-M (n = 12) (top). The same is shown for GPR56-cells for the four groups as a control (bottom). Normalized expression is defined as: (frequency of GPR56+ CD8+ EM cells at D0) * (average IFNG expression within these cells at D1) for each individual. This normalization is performed to assess the combined effects of: 1) increased cell frequency of the GPR56+ cells at baseline (source of IFNγ) and 2) the IFNγ production level after vaccination (D1 IFNG expression) on IFNγ response at D1 in COVR-M. **g**, Scatter plot showing the correlation between GPR56+ CD8 EM cell frequency (as fractions of total CD8 EM in the CITE-seq data; x-axis) and BTM-M7.3 T cell activation signature score of CD8 EM cells computed using CITE-seq pseudobulk gene expression data (y-axis). Spearman correlation and p values are shown. **h**, Related to Fig. 3h but showing CD45RA and CD45RO only with CD8+ TEMRA cells included as an additional comparator. **i**, (left) Circos plot of T-cell receptor (TCR) clonality for different CD8+ T-cell subsets at D0. Segments in the outer circle represent different CD8+ T-cell populations. Segments in inner circle represent male (M) and female (F) for both COVR and HC subjects. Grey lines connect clones sharing identical CDR3 sequences within each individual. Cell subsets are downsampled for visualization (see Methods). (right) Box plot showing Shannon’s entropy index (y-axis) at D0 for each of the indicated CD8+ populations. Significance of differences is determined by two-tailed Wilcoxon test. Shannon’s entropy index evaluates the TCR repertoire diversity for each sample. Higher indices indicate higher diversity (i.e., fewer shared clones shown in Circos plot). EM = effector memory; CM = central memory; TEMRA = EM cells re-expressing CD45RA. **j**, (left) Circos plot of TCR clonality for GPR56+ CD8 EM cells at different timepoints. Segments in the outer circle represent different days in the study (D0, D1, D28). Segments in the inner circle represent males (M) and females (F) for both COVR and HC subjects. Grey lines connect clones sharing identical CDR3 sequences within each sample. Timepoints are downsampled for visualization purposes (see Methods). (right) Box plot showing Shannon’s entropy index (y-axis) of TCR clonality at each of the indicated time points (D0, D1, D28; x-axis) for GPR56+ CD8 EM T-cells (left) and GPR56+ CD8+ TEMRA (right). Significance of differences is determined by two-tailed Wilcoxon test. **k**, (left) Similar to (**j**), but showing the shared clones among different timepoints: segments in the outer circle denote different timepoints. Segments in the inner circle represent unique clones for each individual. Clones and lines connecting shared clones are colored. Samples with less than 30 cells were filtered out for visualization purposes. (right) Line chart showing frequencies of each clone (y-axis) shown in Circos plot (left) at D0, D1 and D28 for each subject. P-values of paired Wilcoxon test are shown comparing the clone frequency differences among D0, D1 and D28.

**Extended Data Figure 5.**
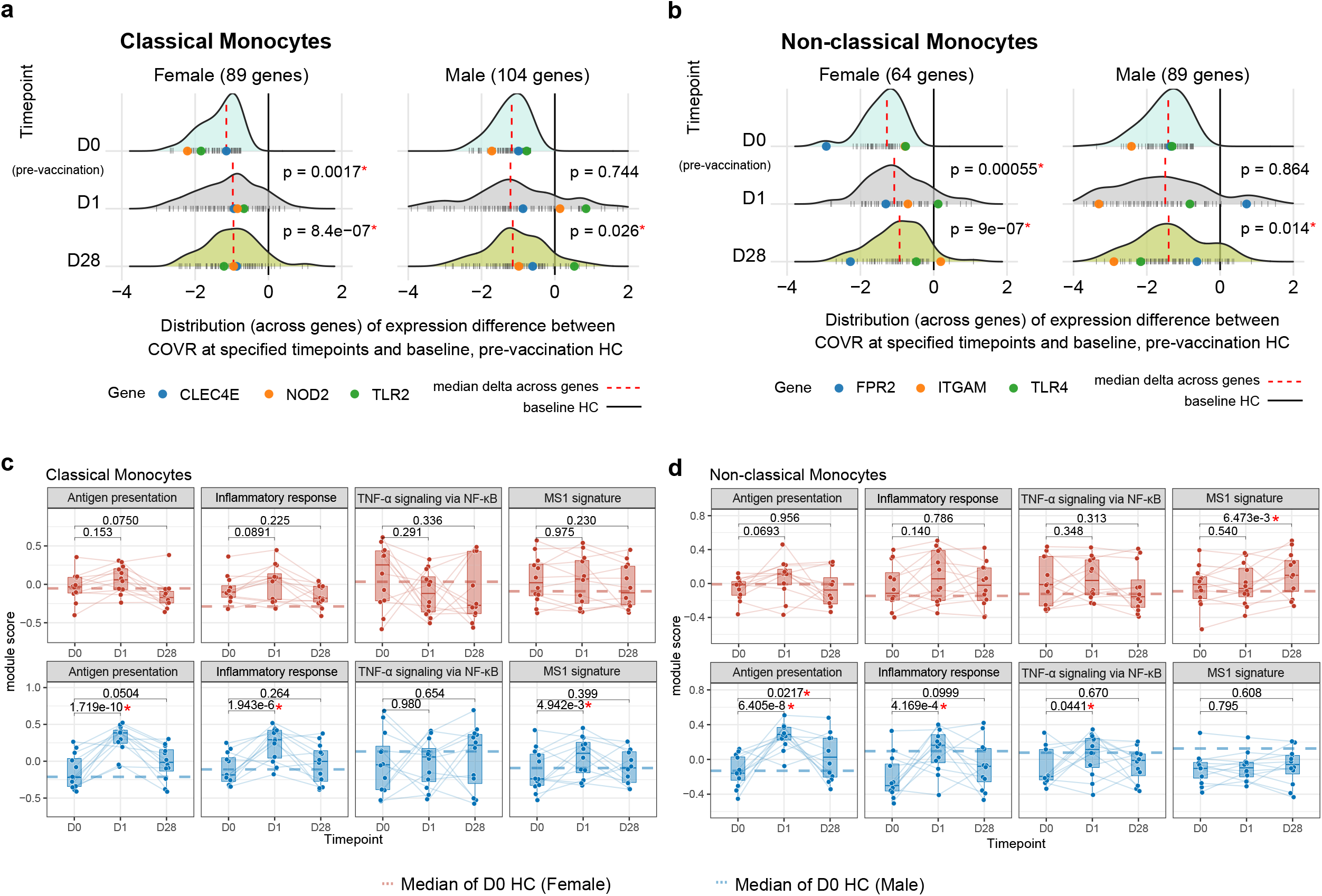
Changes in immune states in COVID-19-recovered individuals following influenza vaccination. **a**, Distributions of gene-level difference of the repressed signature in classical monocytes separately for females (F) and males (M) [shown as z-scores, on a per gene level, capturing the average difference between COVID Recovered (COVR) at the indicated timepoint (top to bottom: D0, D1, and D28) and healthy control (HC) at D0; see Methods]. The gene set comprising the signature includes the union of the BTM-M4.0 and M11.0 leading-edge genes (see Fig. 1f). See also Extended Data Table 5. Dashed red vertical lines represent the median of the distribution. Dark tick marks at the bottom represent individual genes and colored dots highlight specific genes of interest. Significance of differences from D0 is determined by paired two-tailed Wilcoxon test. **b**, Similar to (**a**) but for the non-classical monocytes (with a different gene set comprising the union of the BTM-M4.0 and M11.0 LEGs; see Fig. 1g). **c**, similar to Fig 4b. Boxplots showing the classical monocyte LEG module scores (y-axis) of gene sets from Supplementary Information Fig. 2: antigen presentation related gene sets, Hallmark Inflammatory response, Hallmark TNF-α signaling via NF-κB, and MS-1 signature from Reyes *et al*^22^. LEGs from the first three gene sets were repressed gene sets from Liu *et al*^19^ acute COVID vs. HC GSEA test. **d**, Similar to (**c**), but for non-classical monocytes.

**Supplementary Information Figure 1:**
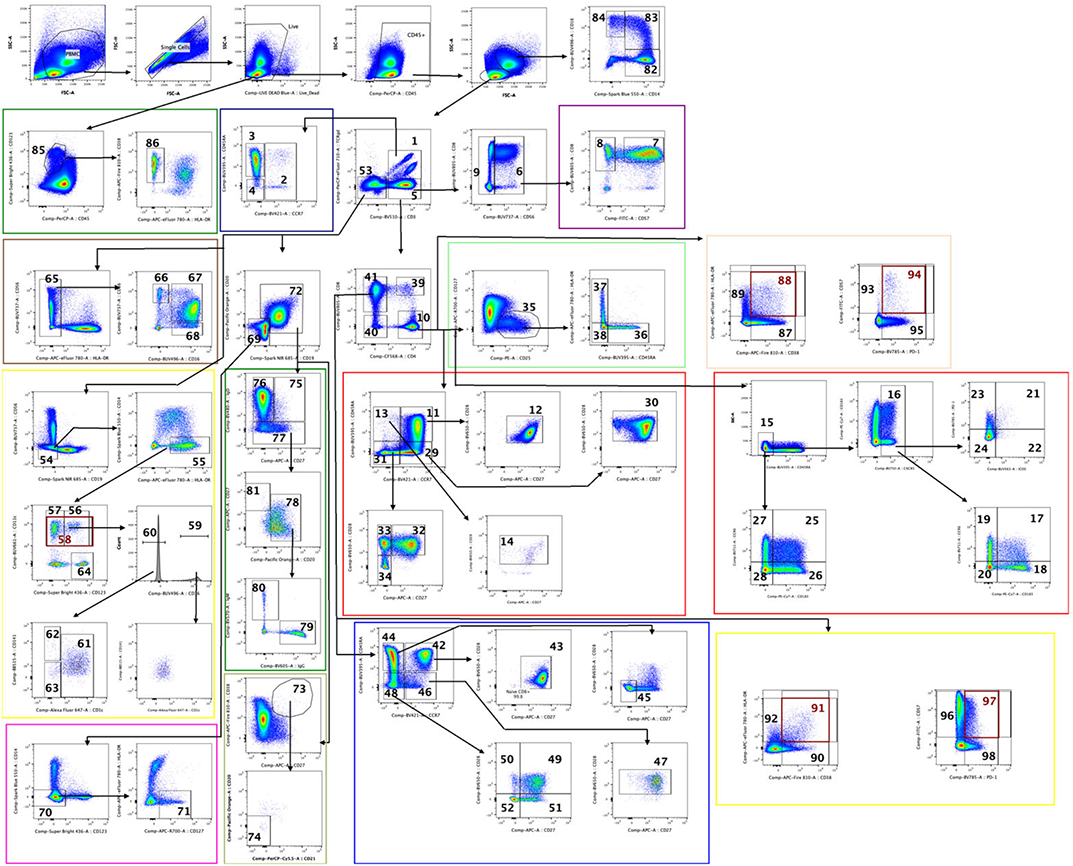
Gating strategy for the Cytek 36-color panel run on peripheral blood mononuclear cells.

**Supplementary Information Figure 2.**
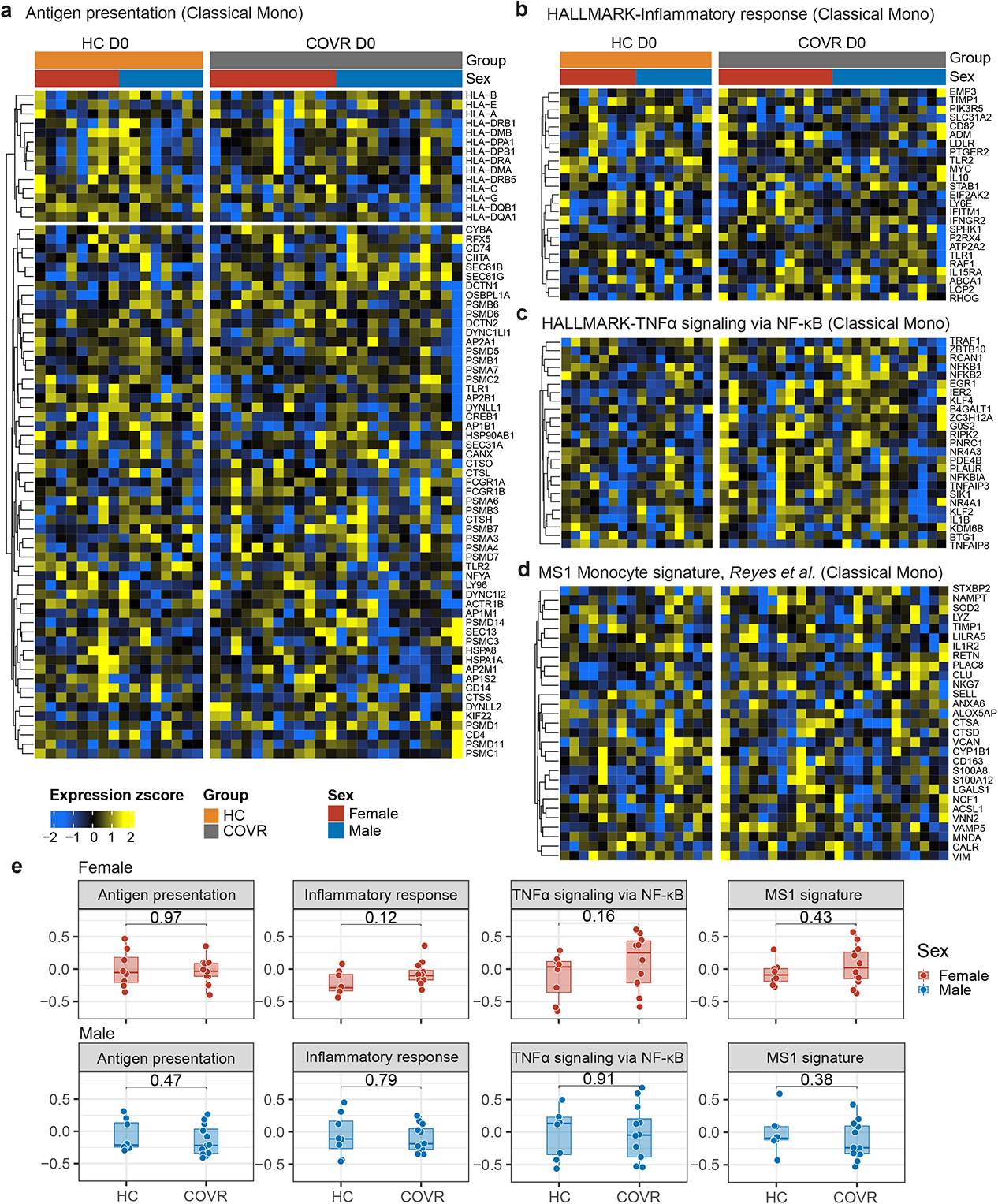
Gene expression profile of antigen presentation, NF-κB/inflammatory, and monocytic myeloid-derived suppressor cell (MDSC) related signatures in classical monocytes. **a,** Heatmap showing the pseudobulk expression of the leading-edge genes (LEGs) from antigen presentation related gene sets, separately for male (M) and female (F), in classical monocytes from the CITE-seq day 0 (D0) pseudobulk data. The LEGs are from the acute COVID-19 vs. healthy control (HC) GSEA analysis in Liu *et al*^19^, which showed that genes in the antigen presentation gene sets – KEGG Antigen processing and presentation, Reactome Antigen processing-Cross presentation, and Reactome MHC class II antigen presentation – tend to be lower in COVID-19. Samples (columns) are grouped by sex and subject group [HC at D0 and COVID-19-recovered (COVR) at D0 as indicated by the bars above the heatmap]. Gene names are shown on the right. **b**, **S**imilar to **(a)**, but showing the LEGs of the “Hallmark Inflammatory response” gene set. **c**, Similar to **(a)**, but showing the LEGs of the “Hallmark TNFα signaling via NF-κB” gene set derived from the acute COVID-19 vs. HC GSEA analysis in Liu *et al*^19^. **d,** Similar to **(a)**, but showing the genes of MSDC/MS1 monocyte signature from Reyes *et al*^22^. **e,** Box plots showing the module scores of the LEGs of the gene sets in (**a-d**) separately for F (top row) and M (bottom row) for the indicated subject groups (columns), in classical monocytes from the CITE-seq D0 pseudobulk data. Each dot represents a sample. P values shown are from two-tailed Wilcoxon tests of the indicated two group comparisons.

**Supplementary Information Figure 3:**
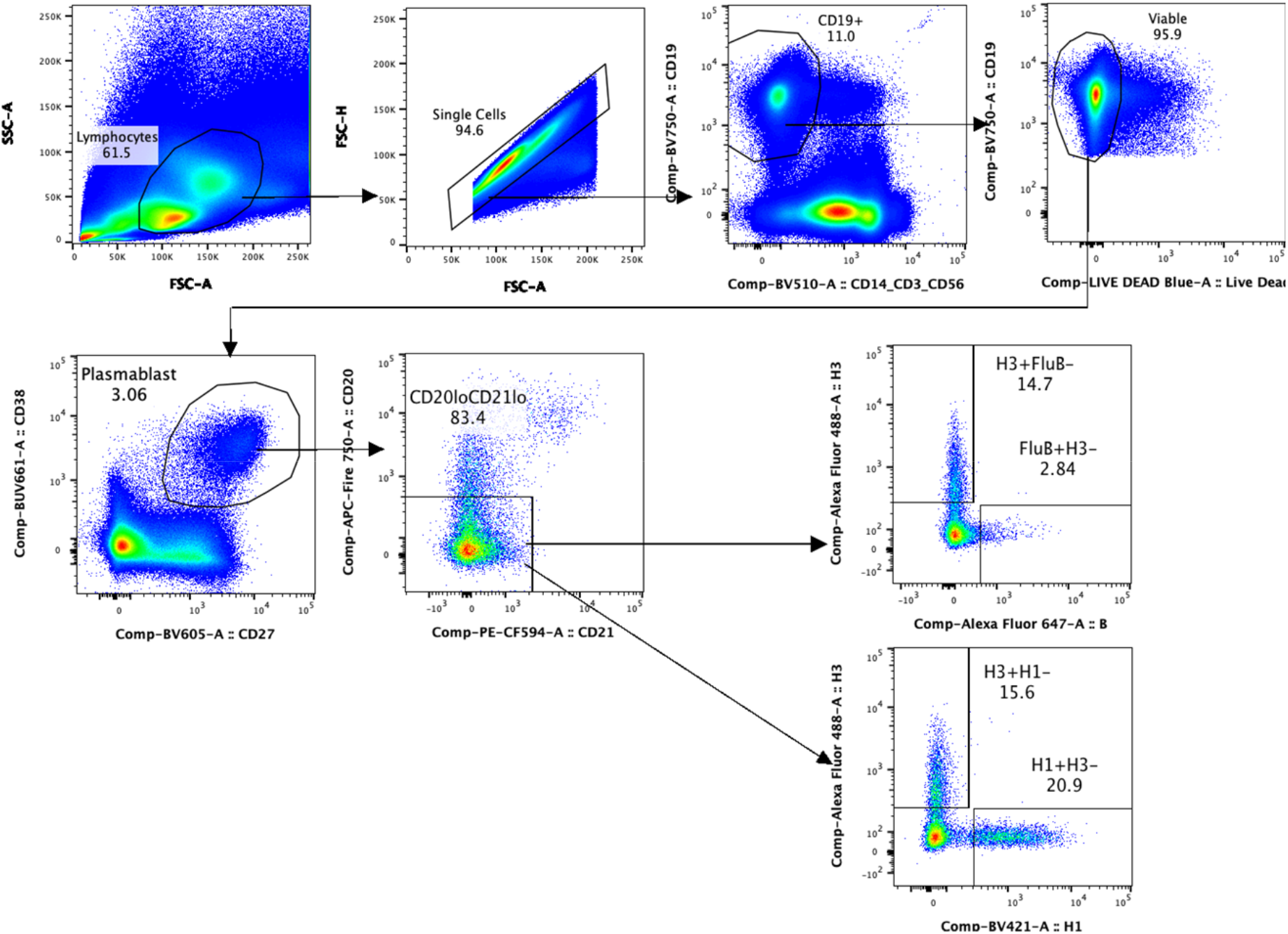
Gating strategies for the influenza-specific plasmablast populations.

## Methods

### Patient population and sample collection

Subjects at least 18 years of age were recruited from the local area (Maryland, Virginia, and the District of Columbia) and enrolled on National Institutes of Health (NIH) protocol 19-I-0126 (Systems analyses of the immune response to the seasonal influenza vaccine). The study was approved by the NIH Institutional Review Board (ClinicalTrials.gov ID: NCT04025580) and complied with all relevant ethical regulations. Informed consent was obtained from all participants. After informed consent, a baseline history and physical examination were performed. Subjects were asked to characterize any present, persistent symptoms of past SARS-CoV-2 infection. Exclusion criteria included obesity (BMI ≥ 30); history of or suspicion of any autoimmune, autoinflammatory or immunodeficiency disease; history of any vaccine within the past 30 days (live attenuated) or 14 days (non-live attenuated); history of any experimental vaccine; history of a parasitic, amebic, fungal, or mycobacterial infection in the past year; or current infection. The COVID-19 vaccine was not available at the time of the study, and no study participants participated in any COVID-19 vaccine trials.

Samples were collected on subjects from three groups: 1) those with a prior history of symptomatic SARS-CoV-2 infection (defined as a history positive nasal PCR test and positive Food and Drug Administration (FDA) Emergency Use Authorization (EUA) SARS-CoV-2 antibody test at the time of protocol screening), 2) those with a history of asymptomatic SARS-CoV-2 infection (defined as a positive FDA EUA SARS-CoV-2 antibody test at the time of protocol exam but no history of COVID-like symptoms; no time since COVID-19 infection or diagnosis (TSD) was identifiable for this group and they were excluded from all TSD analyses), and 3) individuals with no history of SARS-CoV-2 infection (defined as a negative FDA EUA SARS-CoV-2 antibody test at the time of the protocol screening).

Blood for PBMCs, serum, whole blood RNA [Tempus™ Blood RNA Tube (Thermo Fisher Scientific, Waltham, MA)], complete blood count with differential (CBC) and lymphocyte phenotyping was collected at each of the following timepoints relative to seasonal influenza vaccination (day 0): days -7, 0, 1, 7, 14, 28, 70, 100. Optional stool was collected at days 0, 28 and 100. Subjects were provided with Cardinal Health Stool Collection kits (Cardinal Health, Dublin, OH) and Styrofoam storage containers with ice packs to collect stool samples at home and return in person to the NIH. Following day 100, subjects had the option to continue to provide monthly blood samples for PBMCs, serum, whole blood RNA, CBC with differential and lymphocyte phenotyping through August 2021.

At each timepoint following study enrollment, data were collected and managed using REDCap electronic data capture tools hosted at the NIH^1, 2^. REDCap (Research Electronic Data Capture) is a secure, web-based software platform designed to support data capture for research studies, providing 1) an intuitive interface for validated data capture; 2) audit trails for tracking data manipulation and export procedures; 3) automated export procedures for seamless data downloads to common statistical packages; and 4) procedures for data integration and interoperability with external sources. REDCap electronic questionnaires were utilized to collect information from participants via two separate IRB-approved surveys. A survey to evaluate vaccine-related adverse events or symptoms was administered on study days 1 and 7 and a separate survey to evaluate for any health changes or new medications was administered at every visit starting on Day 0. Surveys were sent via email to the participants and responses were transferred from the REDCap system to the NIH Clinical Research Information Management System (CRIMSON) system by the study team.

### Influenza vaccination

Subjects between ages 18 – 64 years were administered the Flucelvax Quadrivalent seasonal influenza vaccine (2020-2021; Seqirus Inc, Summit, NJ). Subjects 65 years of age and older were administered the high-dose Fluzone Quadrivalent seasonal influenza vaccine (2020-2021; Sanofi Pasteur Inc, Swiftwater, PA).

### Influenza microneutralization titers

Virus-neutralizing titers of pre- and post-vaccination sera were determined in a microneutralization assay based on the methods of the pandemic influenza reference laboratories of the Centers for Disease Control and Prevention (CDC) using low pathogenicity vaccine viruses and MDCK cells. The X-179A virus is a 5:3 reassortant vaccine containing the HA, NA, and PB1 genes from A/California/07/2009 (H1N1pdm09) and the 5 other genes from A/PR/8/34 were donated by the high growth virus NYMC X-157. Immune sera were also tested for neutralization titers of the seasonal vaccine strains H1N1 A/Brisbane/59/07, H3N2 A/Uruguay/716/07, and B/Brisbane/60/2001. Internal controls in all assays were sheep sera generated against the corresponding strains at the Center for Biologics Evaluation and Research, FDA, Bethesda, MD. All individual sera were serially diluted (2-fold dilutions starting at 1:10) and were assayed against 100 TCID50 of each strain in duplicates in 96-well plates (1:1 mixtures). The titers represent the highest dilution that completely suppressed virus replication.

### SARS-CoV-2 pseudovirus production and neutralization assay^3–5^

Human codon-optimized cDNA encoding SARS-CoV-2 S glycoprotein (NC_045512) was cloned into eukaryotic cell expression vector pcDNA 3.1 between the *BamH*I and *Xho*I sites. Pseudovirions were produced by co-transfection of Lenti-X 293T cells with psPAX2(gag/pol), pTrip-luc lentiviral vector and pcDNA 3.1 SARS-CoV-2-spike-deltaC19, using Lipofectamine 3000. The supernatants were harvested at 48h post transfection and filtered through 0.45-µm membranes and titrated using 293T-ACE2 cells (HEK293T cells that express ACE2 protein). The following reagent was obtained through BEI Resources, NIAID, NIH: Human Embryonic Kidney Cells (HEK-293T) Expressing Human Angiotensin-Converting Enzyme 2, HEK-293T-hACE2 Cell Line, NR-52511.

For the neutralization assay, 50 µL of SARS-CoV-2 S pseudovirions were pre-incubated with an equal volume of varying dilutions of serum at room temperature for 1 h, then virus-antibody mixtures were added to 293T-ACE2 cells in a 96-well plate. After 3 h incubation, the inoculum was replaced with fresh medium. After 24 hours, cells were lysed and luciferase activity was measured. Controls included cell only control, virus without any antibody control and positive control sera.

### SPR based antibody binding kinetics of human serum^6–8^

Steady-state equilibrium binding of serum was monitored at 25°C using a ProteOn surface plasmon resonance (BioRad). The purified recombinant SARS-CoV-2 or other proteins were captured to a Ni-NTA sensor chip (BioRad, Catalog number: 176-5031) with 200 resonance units (RU) in the test flow channels. The protein density on the chip was optimized such as to measure monovalent interactions independent of the antibody isotype. Serial dilutions (10-, 30- and 90-fold) of freshly prepared sample in BSA-PBST buffer (PBS pH 7.4 buffer with Tween-20 and BSA) were injected at a flow rate of 50 µL/min (120 sec contact duration) for association, and disassociation was performed over a 600-second interval. Responses from the protein surface were corrected for the response from a mock surface and for responses from a buffer-only injection. Total antibody binding was calculated with BioRad ProteOn manager software (version 3.1). All SPR experiments were performed twice, and the researchers performing the assay were blinded to sample identity. In these optimized SPR conditions, the variation for each sample in duplicate SPR runs was <5%. The maximum resonance units (Max RU) data shown in the figures were the RU signal for the 10-fold diluted serum sample.

### PBMC isolation

PBMC samples were isolated from blood collected in Vacutainer EDTA tubes (generic lab supplier) using the SepMate™-50 tubes (STEMCELL Technologies, Cambridge, MA) with following modifications to the manufacturer’s protocol: The blood samples were diluted 1:1 with room temperate PBS and mixed by pipetting. The diluted blood was layered on top of 15ml Cytiva™ Ficoll™ PAQUE-Plus (Cytiva Life Sciences, Marlborough, MA) layer in SepMate™. The SepMate™ tubes were spun at 1200 g for 10 mins with brake set to 5 at room temperature. Following the spin, the top plasma layer was removed as much as possible without disturbing the PBMC layer. If there were any cells stuck on the wall of the tube, then they were gently scraped from the wall with pipette, so they can be resuspended with rest of the cells. The cells were poured from SepMate™ in to a 50ml conical tube. The tubes containing cells were filled up to 50ml with cold wash buffer (PBS with 2% FBS) and mixed by inverting. The tubes were spun at 300 g for 10 mins with brake set to 5 at room temperature. After the spin, the supernatant was removed without disturbing the cell pellet. After resuspending the pellet with cold wash buffer, the cells were counted using the Guava® Muse® Cell Analyzer (Luminex Corporation, Austin, TX). The tubes were again spun at 300 g for 10 mins with brake set to 5 at room temperature. The supernatant was removed without disturbing the cell pellet.

Based on the cell count, 6 – 10 million PBMC were frozen per vial for each sample. Since the cells were counted prior to the last spin, a 50% cell loss was assumed and accounted for in the calculations from cell count. The cell pellet was resuspended with n*600µl (n = number of PBMC vials to be frozen) freezing media (RPMI with 10% FBS) by gentle pipetting. After freezing media, n*600µl DMSO freeze (FBS with 15% DMSO) was added drop-by-drop while gently shaking the tube. In other words, for each vial of PBMC that was to be frozen, 600µl of freezing media and 600µl of DMSO freeze was added, bringing the total volume for each vial to 1.2ml. The solution was gently mixed by pipetting before transferring 1.2ml cell solution to each 1.8ml cryovial (general lab supplier). The cell vials were placed in CoolCell Containers (Thomas Scientific, Swedesboro, NJ) and the container was placed in a −80°C freezer. After at least 4 hours, the PBMC vials were transferred to liquid nitrogen.

### RNA isolation

Blood was drawn directly into the Tempus™ Blood RNA Tube (Thermo Fisher Scientific, Waltham, MA) according to manufacturer’s protocol. Two Tempus tubes were collected at each study timepoint. The blood sample from each Tempus tube was aliquoted in to two 4.5mL cryovials (General lab supplier). These cryovials were directly stored at −80°C.

The RNA samples were isolated in groups of 12-22 samples per batch based on careful batching prior to isolation to reduce confounding factors due to age, gender, and patient group.

RNA was isolated from tempus blood using the QIAsymphony RNA Kit (Qiagen, Gaithersburg, MD) on QIAsymphony SP instrument (Qiagen, Gaithersburg, MD). Blood samples were thawed on ice before each sample was transferred to a 50ml conical tube. The total volume of the sample was brought to 12ml by adding 1x PBS. The tubes were vortexed at full speed for 30 seconds, followed by centrifugation at 3500 g for 1 hour at 4°C. After centrifugation, the supernatant from the tubes was decanted and tubes were placed upside down on clean paper towels for 2 minutes to allow residual liquid to drain. To resuspend the pellet, 800µl of RLT+ buffer was added to the bottom of each tube and vortexed for few seconds. All 800µl of each sample was transferred to 2ml screw cap tubes (Sarstedt, Nümbrecht, Germany). The tubes were placed into #3b adapters (Qiagen, Gaithersburg, MD) to be loaded on to the QIAsymphony.

On the QIAsymphony, RNA CT 800 protocol was selected and used for RNA isolation. The instrument was set up according to the manufacturer’s protocol and the elution volume for RNA samples was set to 100µl. The final volume of the eluted RNA samples ranged from 65 – 95 µl.

RNA yields were determined using Qubit RNA BR kit or Qubit RNA HS kit (Thermo Fisher Scientific, Waltham, MA) based on the yield. RNA RIN numbers were measured using RNA ScreenTape (Agilent Technologies, Santa Clara, CA). The average RIN was 8.3 and average yield was 81.3 ng/µl for the RNA samples.

### RNA-seq

RNA-seq libraries were prepared manually using Universal Plus mRNA-Seq with NuQuant, Human Globin AnyDeplete (Tecan Genomics, Redwood City, CA) according to manufacturer’s protocol. For each sample, 500ng of total RNA was used to isolate mRNA via poly(A) selection. Captured mRNA was washed, fragmented, and primed with the mix of random and oligo(dT) primers. After cDNA synthesis, ends were repaired and ligated with Unique Dual Index (UDI) adaptor pairs. Unwanted abundant transcripts from rRNA, mtRNA and globin were removed using AnyDeplete module. Remaining library was amplified by 14 cycles of PCR and purified with AMPure XP reagent (Beckman Coulter, Indianapolis, IN).

Library concentration was determined by Quant-iT™ PicoGreen™ dsDNA Assay kit (Thermo Fisher Scientific, Waltham, MA) on BioTek Synergy H1 plate reader (BioTek Instruments, Winooski, VT) using 2 ul sample. Library size distribution was determined using D1000 ScreenTape (Agilent Technologies, Santa Clara, CA) on 4200 TapeStation System (Agilent Technologies, Santa Clara, CA). Thirty-two samples were randomly selected from each plate to measure the library size distribution. To determine fragment size, the region on the electropherogram was set from 200 bp to 700 bp. An average of the fragment sizes was used for the rest of libraries to calculate molarity.

To create a balanced pool for sequencing, all libraries from one plate were diluted to the same molar concentration by the QIAgility liquid handling robot (Qiagen, Gaithersburg, MD) and equal volumes of normalized samples were pooled. Ninety-six samples were pooled from each plate on Plates 1-4 and 35 samples were pooled from Plate 5. For an accurate quantification of the pooled libraries, a qPCR was performed using KAPA Library Quantification Kit (Roche, Wilmington, MA).

All libraries were sequenced on the NovaSeq 6000 instrument (Illumina, San Diego, CA) at Center for Cancer Research Sequencing Facility, National Cancer Institute. The libraries pooled from Plates 1-4 were sequenced using one NovaSeq 6000 S4 Reagent Kit (200 cycles) and NovaSeq XP 4-Lane Kit (Illumina, San Diego, CA) with sequencing parameter as 100 bp paired-end reads. The library pool from Plate 5 was sequenced using a NovaSeq 6000 SP Reagent Kit (300 cycles; Illumina, San Diego, CA) with 150 bp paired-end reads as sequencing parameter.

Additionally, after quality control, 11 samples were re-sequenced as Plate 6 on a NextSeq 500 instrument using a NovaSeq 6000 S4 Reagent Kit (200 cycles) with sequencing parameter as 100 bp paired-end reads. Technical replicates were placed on each plate to control for plate variability.

### Serum isolation

Serum was collected directly in Serum Separator Tubes, and allowed to clot at room temperature for a minimum of 30 minutes. Within two hours of blood collection, the tubes were spun at 1800 g for 10 minutes at room temperature. The top (serum) layer was removed via pipette and stored in individual vials at −80°C.

### Complete Blood Counts and lymphocyte phenotyping

Subjects had standard complete blood counts with differential (CBCs) performed at the NIH Clinical Center in the Department of Laboratory Medicine. Lymphocyte (T cell, B cell, NK cell) flow cytometry quantification was performed using the BD FACSCanto™ II flow cytometer (BD Biosciences, Franklin Lakes, NJ).

### Flow cytometry

#### a) B cell panel including influenza HA probes

Thawed PBMC were washed in RPMI culture medium containing 50U/ml benzonase nuclease and then washed by PBS. Cells were incubated with LIVE/DEAD Fixable Blue Dye (Life Technologies, Carlsbad, CA), which was used to exclude dead cells from analysis. Cells were incubated with fluorochrome-conjugated HAs for influenza B (B/Washington/02/2019 and B/Phuket/3073/2013 combined on the same fluorochrome), and Influenza A H1 (A/Hawaii/70/2019) and H3 (A/Hongkong/2671/2019) and fluorochrome-conjugated antibodies against IgM, IgA, CD21, CD85J, FCRL5, CD20, IgG, CD38, CD14, CD56, CD3, CD27, CD71, CD19, IgD for 30 min at 4 C in the dark. The dyes and detailed information of antibodies in the panel (Sarah Andrews, Vaccine Research Center, National Institute of Allergy and Infectious Diseases, National Institutes of Health) are summarized in Table 1. After incubation with antibodies for 30 minutes, cells were washed two times with FACS buffer (0.1%BSA/PBS (pH7.4)) and fixed in 1% paraformaldehyde. Five million cells were acquired on Cytek Aurora spectral cytometer (Cytek Biosciences, Fremont, CA). Data were analyzed with FlowJo software version 10 (BD Biosciences).

**Table 1.**

| Specificity | Fluorochrome | Ab clone | vendor | cat# |
| --- | --- | --- | --- | --- |
| H3 Probe | Alexa Fluor488 |  |  |  |
| IgM | BB700 | G20-127 | BD | Custom |
| IgA | PE | IS11-8E10 | Miltenyi | 130-113-476 |
| CD21 | PE-CF594 | B-Ly6 | BD | 563474 |
| CD85J | PE-Cy7 | GHI/75 | Biolegend | 333712 |
| B Probe | AlexaFluor 647 |  |  |  |
| FCRL5 | R718 | 509F6 | BD | 751885 |
| CD20 | APC-Fire 750 | 2H7 | Biolegend | 302358 |
| IgG | BUV395 | G18-145 | BD | 564229 |
| Dead cells | Live/Dead Blue |  | ThermoFisher | L23105 |
| CD38 | BUV661 | HIT2 | BD | 612969 |
| H1 Probe | BV421 |  |  |  |
| CD14 | BV510 | M5E2 | Biolegend | 301842 |
| CD56 | BV510 | HCD56 | Biolegend | 318340 |
| CD3 | BV510 | OKT3 | Biolegend | 317332 |
| CD27 | BV605 | O323 | Biolegend | 302830 |
| CD71 | BV650 | CY1G4 | Biolegend | 334116 |
| CD19 | BV750 | SJ25C1 | BD | 747161 |
| IgD | BV785 | IA6-2 | BD | 740997 |

#### b) Phenotyping panel

Thawed PBMC were washed in RPMI culture medium containing 50U/ml benzonase nuclease and then washed by PBS. Cells were incubated with LIVE/DEAD Fixable Blue Dye (Life Technologies, Carlsbad, CA), which was used to exclude dead cells from analysis. Cells were washed in FACS staining buffer (1 X phosphate-buffered saline, 0.5% fetal calf serum, 0.5% normal mouse serum, and 0.02% NaN_3_) and incubated with Human Fc block reagent (BD bioscience #564220) at room temperature for 5 min. Cells stained at room temperature for 10 minutes in the dark with fluorochrome-conjugated antibodies against CCR7, CCR6, CXCR5, CXCR3 and TCRgd. Then, stained with fluorochrome-conjugated antibodies against CD45RA, CD16, CD11c, CD56, CD8, CD123, CD161, IgD, CD3, CD20, IgM, IgG, CD28, PD-1, CD141, CD57, CD45, CD25, CD4, CD24, CD95, CD27, CD1c, CD127, HLA-DR, CD38, ICOS, CD21, CD19, CD14 at room temperature for 30 minutes in the dark. Cells were washed two times with FACS staining buffer (1 X phosphate-buffered saline, 0.5% fetal calf serum, 0.5% normal mouse serum, and 0.02% NaN_3_) and fixed in 1% paraformaldehyde. Table 2 shows all the clones and information of antibodies used in the phenotyping panel. A million PBMC were acquired by using Cytek Aurora spectral cytometer (Cytek Biosciences, Fremont, CA). The frequency of major populations was analyzed using with FlowJo™ software version 10 (BD Biosciences) based on previously described manual gating strategies^9–11^.

**Table 2.**

| Specificity | Fluorochrome | Ab clone | vendor | cat# |
| --- | --- | --- | --- | --- |
| CD45RA | BUV395 | 5H9 | BD | 740315 |
| CD16 | BUV496 | 3G8 | BD | 612944 |
| ICOS | BUV563 | DX29 | BD | 741421 |
| CD11c | BUV661 | B-Ly6 | BD | 612967 |
| CD56 | BUV737 | NCAM16.2 | BD | 612766 |
| CD8 | BUV805 | SK1 | BD | 612889 |
| Viability | LIVE/DEAD BLUE |  | Thermo | L34962 |
| CD197 (CCR7) | BV421 | G043H7 | Biolegend | 353208 |
| CD123 | Super Bright 436 | 6H6 | Thermo | 62-1239-42 |
| CD161 | eFluor450 | HP-3G10 | Thermo | 48-1619-42 |
| IgD | BV480 | IA6-2 | BD | 566138 |
| CD3 | BV510 | SK7 | BioLegend | 344828 |
| CD20 | Pacific Organge | HI47 | Thermo | MHCD2030 |
| IgM | BV570 | MHM-88 | Biolegend | 314517 |
| IgG | BV605 | G18-145 | BD | 563246 |
| CD28 | BV650 | CD28.2 | Biolegend | 302946 |
| CD196 (CCR6) | BV711 | G034E3 | Biolegend | 353436 |
| CD185 (CXCR5) | BV750 | RF8B2 | BD | 747111 |
| CD279 (PD-1) | BV785 | EH12.2H7 | Biolegend | 329929 |
| CD141 | BB515 | 1A4 | BD | 566017 |
| CD57 | FITC | HNK-1 | Biolegend | 359604 |
| CD14 | Spark Blue 550 | 63D3 | Biolegend | 367148 |
| CD45 | PerCP | H130 | Thermo | MHCD4531 |
| CD21 | PerCP-Cy5.5 | Bu32 | Biolegend | 354908 |
| TCRgd | PerCP-eFluor710 | B1.1 | Thermo | 46-9959-42 |
| CD25 | PE | BC96 | Thermo | 12-0259-42 |
| CD4 | CF568 | SK3 | Biolegend | Custom |
| CD24 | PE/Dazzle594 | ML5 | Biolegend | 311134 |
| CD95 (Fas) | PE-Cy5 | DX2 | Thermo | 15-0959-42 |
| CD183 (CXCR3) | PE-Cy7 | CEW33D | Thermo | 25-1839-42 |
| CD27 | APC | O323 | Thermo | 17-0279-42 |
| CD1c | Alexa Fluor 647 | L161 | Biolegend | 331510 |
| CD19 | Spark NIR 685 | HIB19 | Biolegend | 302270 |
| CD127 | APC-R700 | HIL-7R-M21 | BD | 565185 |
| HLA-DR | APC/Fire750 | L243 | Thermo | 47-9952-42 |
| CD38 | APC/Fire810 | HIT2 | Biolegend | 303550 |

### Data processing and transformation

#### Bulk RNA-seq data processing

Sequencing reads from Plate 5 were adaptor- and quality-trimmed to 100 bp using Trimmomatic^12^ to match the read length of the other plates (resulting reads with less than 100 bp were discarded). Reads were then aligned to the human genome hg38 using the STAR aligner. Duplicate reads from PCR amplification were removed based on Unique Molecular Identifiers (UMI). Gene expression quantification was performed using the featureCounts^13^ function from Subread package. Samples with less than 5 million assigned reads were re-sequenced and replaced. Reads were normalized and log transformed using *limma voom*^14^. Lowly expressed genes, defined as having fewer than five samples with > 0.5 counts per million reads, were removed. Pre-vaccination (days -7 and 0) samples from the same healthy control (HC) subjects were considered as replicates and were used to estimate latent technical factors by the RUVs function from the SeqRUV^15^ R package. Four latent variables were included to derive normalized gene expression values used for visualization and when specifically noted. Variable genes based on intra-subject variability of pre-vaccination samples in the HCs and across technical replicates were filtered out, resulting in a total of 10017 remaining genes for downstream analyses.

#### OLINK serum proteomics

Missing values were imputed using k-nearest neighbors approach with k=10. For each sample, probes targeting the same protein were averaged.

#### Cytek flow cytometry

Cell frequencies were generated by converting cell counts as fraction of live cells or lymphocytes as specified. The frequency data were log2 transformed for linear modeling. For populations with zero counts in any of the samples, an offset equaling to half of the smallest non-zero value was added across samples.

#### CBC with diff and TBNK

Both absolute and relative counts were log2 transformed for linear modeling. For parameters with zero values in any of the samples, an offset equaling to half of the smallest non-zero value was added across samples.

#### Baseline differential expression analysis

Using the dream^16^ function in the variancePartition R package, mixed-effects models were applied to determine differential levels of analytes (i.e. whole-blood gene expression, serum proteins, cell frequencies, flu titer and SPR, and hematological parameters) between COVID-recovered and HC subjects in a sex-specific manner as follows:

∼ 0 + group:sex + age + race + batch.effects + (1|subject.id)

Batch effect-related covariates were added to specific models depending on the assay type. For bulk RNA-seq, these include the four latent technical factors (see ***Bulk RNA-seq data processing***) and the timepoint-matched % neutrophils parameter from the CBC panel. For the Cytek and Olink platforms, sampling batch/plate was included as covariates. In addition to day 0, available samples from day -7 (in RNA-seq and CBC panel), were included as baseline replicates in the modeling.

Sex-specific group differences were computed from the contrasts covid.Female – healthy.Female and covid.Male – healthy.Male. Overall COVID vs. HC difference was determined by combining the two contrasts, i.e. (covid.Female – healthy.Female)/2 + (covid.Male – healthy.Male)/2. Sex difference linked to SARS-CoV2 infection was derived from the contrast (covid.male – covid.female) – (healthy.male – healthy.female) to account for normal differences between males and females. P values were adjusted for multiple testing within each assay type and contrast combination using the Benjamini-Hochberg (BH) method (Benjamini and Hochberg, 1995).

#### Association with time since COVID-19 diagnosis

To evaluate whether any of the differences detected at baseline had stabilized or might still be resolving, a linear model was used to test the association of relevant parameters with the time since COVID-19 diagnosis (TSD) among COVID-recovered subjects:

∼ 0 + sex + sex:scale(TSD) + age + race + (1|subject.id)

Two asymptomatic subjects without known TSD were excluded from the model. Association was assessed separately for females and males, and jointly by the combined contrast (Female:TSD + Male:TSD)/2. Dependent variables were converted to ranks in the model to reduce the effect of potential outliers.

Using a conservative approach, genes were classified as TSD-associated if they had an unadjusted p value < 0.05 and were excluded from subsequent analyses as specified. To determine whether the any of the baseline differential gene sets were associated with TSD (e.g. Extended Data Fig. 1h), leading edge gene (LEG) modules were derived from the union of all LEGs of the same gene set from different contrasts (see ***Gene set module scores***). A gene set was considered stable if none of three contrasts tested in the association model were significant (using unadjusted p value threshold of 0.05).

#### Post-vaccination differential expression analysis

Similar to the workflow employed in baseline differential expression analysis, mixed-effects models were created to evaluate changes and group differences at each available timepoint after vaccination. Subjects aged 65 and above were excluded as they received a different type of vaccine. In addition to the baseline covariates, the model also accounts for the participants’ flu vaccination history within last 10 years as follows:

∼ 0 + visit:group:sex + age + race + flu.vax.count.10yr + batch.effects + (1|subject.id)

Three types of comparisons were examined using this model:

**1. Timepoint-specific group differences** Similar to the contrasts in the baseline model, but for individual timepoints post vaccination (day 1 to day 100).
**2. Vaccine-induced changes in group difference** Similar to the timepoint-specific contrasts above, but additionally subtracting off the corresponding baseline contrast to assess changes relative to the baseline. For example, vaccine-induced changes for female COVID vs. HC differences at D1 is evaluated with the contrast: (D1.covid.Female – D1.healthy.Female) – (Baseline.covid.Female – Baseline.healthy.Female).
**3. Reversal of COVID vs. HC difference** Instead of using the HC subjects at the same corresponding timepoints as reference, post-vaccination samples from the COVID-recovered subjects were compared to baseline HC with the contrasts [timepoint].covid.Female – baseline.healthy.Female and [timepoint].covid.Male – baseline.healthy.Male. These contrasts can inform whether any pre-vaccination differences observed in the COVID-recovered subjects were reverted towards healthy baseline levels after vaccination. Reversal is defined as having smaller absolute effect size (using the z.std value from the dream function) at D1 and D28 after vaccination compared to the baseline absolute effect size.

P values were adjusted for multiple testing per each timepoint, assay type and contrast combination using the BH method.

#### Gene set enrichment of differentially expressed (DE) genes

Enriched gene sets were identified using the pre-ranked gene-set enrichment analysis (GSEA) algorithm implemented in the clusterProfiler R package^17^. Genes were ranked using signed −log10 p values from differential expression models. Enrichment was assessed with gene set lists from MSigDB’s Hallmark collection^18^, Blood Transcriptomic Modules^19^, and cell type gene signatures^20^. Only gene sets with 10 to 300 genes were considered. P values were adjusted per gene set list for each contrast using the BH method and gene sets with FDR < 0.05 were considered significant. Baseline enriched gene sets were derived by intersecting significant gene sets extracted from DE models using samples independently from day -7, day 0, and both days combined. Genes associated with time since diagnosis (TSD) at baseline (see ***Association with time since COVID-19 diagnosis***; Extended Data Table 3) were excluded from the post-vaccination enrichment analyses to help segregate the effect of vaccination from natural temporal resolution of the SARS-CoV-2 infection.

#### Gene set module scores

Gene set module scores were generated from SeqRUV normalized gene expression values (see ***Bulk RNA-seq data processing and transformation***) using gene set variation analysis (GSVA) method in GSVA R package^21^. LEG module scores representing enriched pathway activities were calculated for relevant samples using LEGs identified by GSEA to enhance signal-to-noise ratio. The average scores between days -7 and 0 were used for calculating post-vaccination changes relative to baseline.

#### Endpoint association

To evaluate the association of relevant parameters, including gene set module scores and cell frequencies, with interferon (IFN) or antibody titer fold change endpoints, the following model was applied:

endpoint ∼ group:sex + scale(parameter):group:sex + age + race + flu.vax.count.10yr

The endpoint values were converted to rank to reduce the effects of potential outliers. Replicates from the same subjects were averaged.

#### Serology

Influenza antibody titers below the detection limit of 1:20 were set to 1:10. Maximum titer across strains was calculated by normalizing titer levels across all samples from both day 0 and day 28 individually for each of the four strains followed by taking the maximum standardized titer for each sample.

#### Baseline titer difference analysis

For each of the four strains, a linear model was applied to determine baseline titer differences between COVID-recovered and HC subjects in a sex-specific manner as follows:

day 0 titer ∼ group:sex + age + race

Titer values were log10 transformed in the model, and sex-specific group differences were computed from the contrasts covid.Female – healthy.Female and covid.Male – healthy.Male. Subjects aged 65 and above were excluded from the analysis.

#### Day 28 titer difference analysis

For post-vaccination titer response, both day 28 titer and day 28/day 0 fold change (FC) were evaluated as endpoints to determine differences between COVID-recovered and HC subjects for each of the four strains:

endpoint ∼ group:sex + age + race + flu.vax.count.10yr + day 0 titer

For day 28 FC, a negative binomial model with log link was applied using glm.nb function in the MASS R package. A linear model was used to fit the day 28 titers. Strain-specific titer values were log10 transformed in the model. Group differences were assessed using the same subjects and contrasts as in the baseline analysis.

Influenza antibody avidity as measured by surface plasmon resonance (SPR) were analyzed in the same manner as the titer data across HA1 and HA2, with the exception that that a linear model was applied for the fold changes.

#### Concordance in natural influenza infection cohort

A prospective cohort study with subjects profiled prior to and at least 21 days after natural influenza infection in two seasons^22^ was utilized to assess residual effects of the infection separately in males and females. Gene expression data were downloaded from GEO using the accession GSE68310. Subjects with only influenza A virus infection (n=51 females and 35 males) were identified and included for this analysis. Lowly expressed probes were removed, and the remaining data were converted to gene-based expressions. No additional processing steps were performed as the data were already normalized.

Separately for each season, differential expression analysis between baseline (pre-infection) and spring (long term post-infection) samples from the same individuals were performed using the dream function in the variancePartition R package. A mixed-effects model accounting for flu vaccination history and disease severity (based on fever grade: none, low, and high) was constructed as follows:

∼ 0 + timepoint:sex + age + num.flu.vaccination + fever.grade + (1|subject.id)

Differentially expressed (DE) genes were identified using the contrasts Spring.F - Baseline.F and Spring.M – Baseline.M for females and males, respectively. Sex difference was evaluated by the contrast (Spring.M – Baseline.M) – (Spring.F - Baseline.F). Concordance of DE results between the two seasons were evaluated based on correlation of effect size across genes (z.std values generated by dream).

Enrichment analysis was performed to determine whether the same set of genes were differentially expressed between pre- and post-influenza infection from this independent cohort and in COVID-recovered subjects compared to healthy controls prior to vaccination. To better match the age range of subjects between the two studies, baseline differential gene analysis was performed again with subjects under 65 years of age in the COVID cohort (see ***Baseline differential expression analysis***). Given that the males showed stronger concordance between the two flu seasons (Extended Data Fig. 2b), COVID DE genes were ranked by signed −log10 p values and tested against a gene set formed by the intersect of DE (p < 0.05) genes in males from the flu infection cohort.

### CITE-seq

#### a) Single cell CITE-seq processing

Frozen PBMC samples were thawed, recovered and washed using RPMI media with 10% FBS and 10mg/mL DNase I (STEMCELL) and then processed as previously described^23^ for CITE-seq staining. In brief, samples from different donors were pooled and different timepoints from the same donor were pooled separately so that each pool contains only one timepoint from one donor. PBMC pools were Fc blocked (Human TruStain FcX, BioLegend) and stained with Totalseq-C human ‘hashtag’ antibodies (BioLegend), washed with CITE-seq staining buffer (2% BSA in PBS). Then hashtagged PBMC pools were combined and cells were stained with a cocktail of TotalSeq-C human lyophilized panel (BioLegend) of 137 surface proteins (including 7 isotype controls, refer to repository table) and SARS-CoV-2 S1 protein probe. Then, cells were washed, resuspended in PBS, and counted before proceeding immediately to the single cell partition step.

#### b) Single cell CITE-seq library construction and sequencing

PBMC samples were partitioned into single cell Gel-Bead in Emulsion (GEM) mixed together with the reverse transcription (RT) mix using 10x 5’ Chromium Single Cell Immune Profiling Next GEM v2 chemistry (10x Genomics, Pleasanton, CA), as previously described^23^. The RT step was conducted in the Veriti™ Thermal Cycler (ThermoFisher Scientific, Waltham, MA). Single cell gene expression, cell surface protein, T cell receptor (TCR) and B cell receptor (BCR) libraries were prepared as instructed by 10x Genomics user guides (https://www.10xgenomics.com/resources/user-guides/). All libraries were quality controlled using Bioanalyzer (Agilent, Santa Clara, CA) and quantified using Qubit Fluorometric (ThermoFisher). 10x Genomics 5’ Single cell gene expression, cell surface protein tag, TCR and BCR libraries were pooled and sequenced on Illumina NovaSeq platform (Illumina, San Diego, CA) using the following sequencing parameters: read1-100-cycle, i7-10-, i5-10, read2-100.

### CITE-seq data processing and statistical analyses

#### a) Single cell sample demultiplexing and preprocessing

Single cell sequencing data was demultiplexed, converted to FASTQ format, mapped to human hg19 reference genome and counted using *CellRanger* (10x Genomics) pipeline. The sample level demultiplex was done based on two levels as previously described^23^: 1) Hashtag antibody staining to distinguish different timepoint samples from a same subject; 2) single nucleotide polymorphisms (SNPs) called from the whole blood RNA-seq data to identify different subjects. Specifically, *CellRanger* version 6.0.1 was used for generating count matrix and the software package *demuxlet* (v2, from the *‘popscle’* software suite)^24^ was used to match single cell gene expression data to each donor and identify empty droplets and doublets.

#### b) Single-cell data clustering and cell annotation

Single-cell data were further processed using Seurat (v4.0.3) running in R v4.1.1. We removed cells with less than 200 and greater than 5,000 detected genes, greater than 60% of reads mapped to a single gene, greater than 15% mitochondrial reads, cell surface protein tag greater than 20,000, and hashtag antibody counts greater than 20,000. The protein data was normalized and denoised using the DSB method^25^. The following parameters were used in the dsb normalization function: define.pseudocount = TRUE, pseudocount.use = 10, denoise_counts = TRUE, use.isotype.control = TRUE. The DSB-normalized protein data were used to generate the top variable features (n = 100) and principal components (PCs). Then the shared nearest neighbor (SNN) graph followed by k-nearest neighbors clustering were built using the FindNeighbors and FindClusters functions using first 15 PCs in Seurat (v4.0.3), respectively. Cell clusters were quality controlled based on their nearest neighbors and cell surface proteins. Cells were then further clustered within each major cell population using “weighted-nearest neighbor” (WNN) analysis in Seurat^26^ (v4.0.3) by integrating both cell surface protein and gene expression modalities. WNN FindMultiModalNeighbors were done using both top 10 PCs for cell surface protein and RNA of variable features. The WNN clusters were manually annotated and QC using the surface protein together with gene expression.

#### c) Effector memory CD8 cell (CD8-EM) annotation for CITE-seq clusters

All CD8 cells were clustered using WNN as described above. CD8 clusters were annotated based on their surface markers as reported^27^ together with gene expression profile. RNA expression of CD8 cells was mapped to external dataset using Seurat Label transfer method^26, 28^ (v4.0.3). Clusters annotated as CD8-EM are surface CD45ROhi, CD45RAlo, CD95+, CD62Llo and CCR7-(mRNA) with most cells (∼90%) mapped to CD8-EM phenotype cells in an external dataset^26, 28^.

#### d) Pseudobulk differential expression and gene set enrichment analysis

Cells from a given sample were computationally “pooled” according to their cell type assignment by summing all reads for a given gene. Pseudobulk libraries made up by few cells and therefore likely not modeled properly by bulk differential expression methods were removed from analysis for each cell-type to remove samples that contained fewer than 4 cells and less than 35000 library size after pooling. Lowly expressed genes were removed for each cell type individually using the filterByExpr function from *edgeR*^29^ with min.count = 2. Log counts per million (cpm) of each gene were calculated with scaling factors for library size normalization provided by the calcNormFactors function. Differential expression analysis was performed using the same models described in ***Post-vaccination differential expression analysis*** without running baseline models separately because the entire CITE-seq cohort was under 65 years of age. Batch assignment and number of barcodes/cells per sample were included as batch effects in this model.

Similarly, gene set enrichment analysis was carried out for each cell type in the same manner as for the bulk RNA-seq data (see ***Gene set enrichment of differentially expressed (DE) genes***) which particular focus on the baseline enriched genesets identified by the bulk RNA-seq analysis. The Monaco gene sets were excluded from the single-cell analysis given the cell clusters were annotated and no further cell type demultiplex needed.

#### e) Gene set module score calculation

Module scores (gene set signature score) representing enriched pathway activities were calculated for each pseudobulk sample of certain cell types. The pseudobulk gene counts were corrected with removeBatchEffect function in *limma* package to remove experimental batch and cell number effects and then normalized with voom^30^. The scores were then generated using gene set variation analysis (GSVA) method from the *GSVA* R package^21^. Specifically, for monocyte signatures, LEGs of BTM modules M4.0 and M11.0 were identified by GSEA from 1) D0.COVR-F vs. D0.HC-F and 2) D0.COVR-M vs. D0.HC-M models. The union of LEGs were used for the score calculation for female and male samples, respectively.

For BTM-M7.3 T cell activation signature and other signatures from acute COVID data as indicated in the figures, LEGs were used from the indicated comparison groups for the score calculation of female and male separately.

For the HALLMARK IFNγ response module score, all genes from the geneset were used for calculation of module scores in each celltype, so that the differences between celltypes can be compared.

#### f) Single-cell module score calculation and Visualization

To visualize the differences between different patient groups in single data of the certain signatures, the genes from indicated genesets were used to calculate the module scores of each single cell. Module scores were calculated using AddModuleScore function in *Seurat* and then visualized in UMAP plots. For D1 vs. D0 HALLMARK IFNg response module score differences shown in umaps, cells from D1.HC-F, D1.COVR-F, D1.HC-M and D1.COVR-M groups were downsampled to the same number of cells. The UMAP embeddings of cells colored with average differences for each high resolution cell subsets are shown.

#### g) Single-cell module score calculation and test of external acute COVID-19 single-cell CITE-seq data

Single-cell data from the Brescia cohort of Liu *et al*^23^ was downloaded from GEO. Single monocytes data was extracted and Single-cell data from the Brescia cohort were pooled as described in “c) Pseudobulk differential expression and gene set enrichment analysis”. The gene set module scores of BTM modules M4.0 and M11.0 for all samples were generated using the union LEGs of male and female in “d) Gene set module score calculation”. The pseudobulk gene counts were normalized with the varianceStabilizingTransformation function from *DEseq2* R package^31^. The scores were then generated using gene set variation analysis (GSVA) method from the *GSVA* R package^21^. Given there are multiple samples from each subject, the differences between patient groups (HC, less severe and more severe, corresponding to HC, DSM-low and DSM-high in Liu *et al*) were tested using the *Limma* linear model, where samples from the same donors were treated as duplicates using *duplicateCorrelation*. P-values of t statistics from the linear model of indicated contrasts are shown.

#### h) Single cell TCR data processing

*CellRanger* version 6.0.1 was used to assemble V(D)J contigs. The V(D)J assignment and clonotype were from the CellRanger output of the filtered contig_annotations.csv file for each 10x lane. The data is combined for all lanes and paired TCRα and TCRβ chains for each single cell were combined using *scRepertoire* R package^32^ and integrated with the single-cell CITE-seq Seurat object metadata. Cells annotated as CD8 T cells and with both α and β chains detected are filtered and analyzed. CD8 subsets and GPR56+ CD8 effector memory cell clonality were visualized with Circos plots using *Circlize* R package^33^. For visualization purpose, cells from each subset were downsampled with equal number in each subset (for comparison between subsets, Extended Data Fig. 4i) or in each timepoint (for comparison between timepoints, Extended Data Fig. 4j,k). Cells were considered as the same clone with identical CDR3 (both α and β chains). Identical clones were connected within each sample or each subject across timepoints with lines.

#### i) TCR diversity metric calculation

Shannon’s entropy (H’ index) was calculated as a measure of TCR diversity^34, 35^. Samples for each CD8 subsets with fewer than 50 cells were filtered from the calculation. All samples were downsampled to 50 cells because the diversity metric can be affected by the sample cell numbers. The process was repeated 1000 times with random downsampling of 50 cells and the median Shannon’s index was used as an estimate of diversity for a given sample. Differences of the diversity metric between different CD8 subsets or timepoints were tested using two-tailed Wilcoxon test.

#### j) Reversal genes and bootstrapping to infer significance of difference in reversal of monocyte repressed signature between COVR-F and COVR-M

Reversal genes are defined as those whose COVR vs. D0 HC absolute effect size (z.std values from dream; see ***Post-vaccination differential expression analysis***) are smaller at both D1 and D28 compared to D0.

Bootstrapping was employed to determine the significance of difference between COVR-F and COVR-M in their proportion of baseline LEGs from the monocyte depressed signature (BTM M4.0 and M11.0) that moved towards baseline HC. Subjects from each subject group were randomly sampled with replacement in each round of the bootstrapping and their samples were analyzed as described in ***Post-vaccination differential expression analysis***. The proportion of LEGs reversed after vaccination were calculated in each round for COVR-F and COVR-M in classical and non-classical monocytes, separately, and the p values plotted in Fig. 4e were determined based on 20 rounds of this procedure.

#### j) Visualization of gene expression in heatmaps

Heatmaps showing pseudo-bulk data was generated using *ComplexHeatmap* R package^36^. The log(CPM) (counts per million) normalized expression for each sample for a given celltype was calculated by pooling cells as described in “c) Pseudobulk differential expression and gene set enrichment analysis”. Heatmaps are showing z-score of the normalized expression for each gene in each sample.

