## Extended Data Table 1 for "Influenza vaccination and single cell multiomics reveal sex dimorphic immune imprints of prior mild COVID-19"

**Extended Data Table 1.** Cohort Characteristics

| | COVR | | Healthy | |
|---|---|---|---|---|
| | **Female** | **Male** | **Female** | **Male** |
| Subject count | 17 | 16 | 21 | 19 |
| **Age (Years)** | | | | |
| Median | 40.2 | 43.7 | 52.5 | 47.6 |
| Mean | 44.9 | 43.7 | 47.2 | 47.4 |
| Min | 23.4 | 21.9 | 22.5 | 24.0 |
| Max | 70.5 | 67.3 | 70.4 | 69.1 |
| Aged > 65 | 2 | 2 | 5 | 5 |
| **Race** | | | | |
| Asian | 1 | 0 | 2 | 2 |
| Black | 1 | 0 | 4 | 0 |
| Multiple race | 0 | 2 | 1 | 3 |
| White | 15 | 14 | 14 | 14 |
| **Number of Influenza Vaccination in Past 10 Years** | | | | |
| 0 | 0 (0%) | 1 (6.25%) | 0 (0%) | 2 (10.53%) |
| 1 | 1 (5.88%) | 0 (0%) | 1 (4.76%) | 0 (0%) |
| 2 | 0 (0%) | 1 (6.25%) | 2 (9.52%) | 2 (10.53%) |
| 3 | 1 (5.88%) | 0 (0%) | 1 (4.76%) | 0 (0%) |
| 4 | 0 (0%) | 0 (0%) | 2 (9.52%) | 0 (0%) |
| 5 | 3 (17.65%) | 3 (18.75%) | 1 (4.76%) | 0 (0%) |
| 6 | 1 (5.88%) | 1 (6.25%) | 1 (4.76%) | 0 (0%) |
| 7 | 3 (17.65%) | 0 (0%) | 1 (4.76%) | 1 (5.26%) |
| 8 | 0 (0%) | 1 (6.25%) | 1 (4.76%) | 3 (15.79%) |
| 9 | 1 (5.88%) | 1 (6.25%) | 1 (4.76%) | 1 (5.26%) |
| 10 | 7 (41.18%) | 8 (50%) | 10 (47.62%) | 10 (52.63%) |
| Experienced side effects after vaccination | 16 (94.1%) | 9 (56.3%) | 17 (90.0%) | 15 (78.9%) |
| **COVID-19 Symptoms** | | | | |
| Asymptomatic | 1 (5.9%) | 1 (6.3%) | - | - |
| Symptomatic | 16 (94.1%) | 15 (93.8%) | | |
| **Time since COVID-19 Diagnosis (Days) *** | | | | |
| Median | 172.0 | 186.0 | - | - |
| Mean | 152.9 | 149.3 | - | - |
| Min | 58.0 | 44.0 | - | - |
| Max | 237.0 | 248.0 | - | - |
| **Duration of acute COVID-19 symptoms (Days) *** | | | | |
| Median | 14 | 10 | | |
| Mean | 19.62 | 13.07 | | |
| Min | 4 | 1 | | |
| Max | 87 | 33 | | |

| | | | | |
|---|---|---|---|---|
| Experienced COVID-19 residual symptoms at time of screening | 8 (47.1%) | 3 (18.8%) | - | - |
| **COVID-19 Residual Symptoms** | | | | |
| Brain fog | 1 (5.9%) | 0 (0%) | - | - |
| Fatigue | 2 (11.8%) | 0 (0%) | - | - |
| Loose stools | 0 (0%) | 1 (6.3%) | - | - |
| Reduced sense of taste | 1 (5.9%) | 1 (6.3%) | - | - |
| Reduced sense of smell / smell disturbances | 5 (29.4%) | 2 (12.5%) | - | - |
| Shortness of breath / Chest pressure | 1 (5.9%) | 0 (0%) | - | - |

* Excluding asymptomatic subjects
